# Disease-dependent interaction policies to support health and economic outcomes during the COVID-19 epidemic

**DOI:** 10.1101/2020.08.24.20180752

**Authors:** Guanlin Li, Shashwat Shivam, Michael E. Hochberg, Yorai Wardi, Joshua S. Weitz

**Affiliations:** Interdisciplinary Graduate Program in Quantitative Biosciences, Georgia Institute of Technology, Atlanta, GA, USA; School of Physics, Georgia Institute of Technology, Atlanta, GA, USA; School of Electrical and Computer Engineering, Georgia Institute of Technology, Atlanta, GA, USA; ISEM, Université de Montpellier, CNRS, IRD, EPHE, Montpellier, France; Santa Fe Institute, Santa Fe, NM 87501, USA; School of Biological Sciences, Georgia Institute of Technology, Atlanta, GA, USA

**Author notes:** Correspondence to: Michael E Hochberg, Yorai Wardi and Joshua S. Weitz. Authors contributed equally to the work (Guanlin Li, Shashwat Shivam).

## Abstract

Lockdowns and stay-at-home orders have partially mitigated the spread of Covid-19. However, the indiscriminate nature of mitigation — applying to all individuals irrespective of disease status — has come with substantial socioeconomic costs. Here, we explore how to leverage the increasing reliability and scale of both molecular and serological tests to balance transmission risks with economic costs involved in responding to Covid-19 epidemics. First, we introduce an optimal control approach that identifies personalized interaction rates according to an individual’s test status; such that infected individuals isolate, recovered individuals can elevate their interactions, and activity of susceptible individuals varies over time. Critically, the extent to which susceptible individuals can return to work depends strongly on isolation efficiency. As we show, optimal control policies can yield mitigation policies with similar infection rates to total shutdown but lower socioeconomic costs. However, optimal control policies can be fragile given mis-specification of parameters or mis-estimation of the current disease state. Hence, we leverage insights from the optimal control solutions and propose a feedback control approach based on monitoring of the epidemic state. We utilize genetic algorithms to identify a ‘switching’ policy such that susceptible individuals (both PCR and serological test negative) return to work after lockdowns insofar as recovered fraction is much higher than the circulating infected prevalence. This feedback control policy exhibits similar performance results to optimal control, but with greater robustness to uncertainty. Overall, our analysis shows that test-driven improvements in isolation efficiency of infectious individuals can inform disease-dependent interaction policies that mitigate transmission while enhancing the return of individuals to pre-pandemic economic activity.

## Introduction

As of 13 August 2020, more than 20,439,000 cases of coronavirus disease 2019 (COVID-19) have been reported worldwide with more than 744,000 deaths globally^1^. Starting at the reported origin of the pandemic in Wuhan, China, control measures have been implemented in most countries where outbreaks have occurred ^2,3,4,5,6,7^. Multiple public health strategies are being deployed to slow outbreaks, and although recommendations always include social distancing and isolation of confirmed cases, the full spectrum of measures and levels of adherence differ from country to country, making assessments of strategy efficacy difficult (see ^2^, controversy surrounding ^8^)

The non-pharmaceutical control strategies for COVID-19 largely follow those employed in previous viral epidemics, including SARS, Ebola and MERS. Initial strategies can be broadly grouped into mitigation and suppression, where the former attempts to preserve essential health care services and contain morbidity and mortality, whereas the latter imposes more severe, emergency restrictions to prevent health care system collapse and provide conditions for easing-off towards less intense mitigation strategies ^9^. Both mitigation and suppression approaches carry considerable social and economic costs, meaning that policymakers and the public at large only adopt them for short time periods ^10^. A problem is that control measures have often been applied irrespective of an individual’s disease status (and/or likely infection risk severity) and are driven, in part, by the absence of information-driven alternatives.

Hence, distinct from lockdowns, there is an increasing interest in implementing population-wide prevention methods that decrease transmission risk while enabling economic re-engagement. Examples of such measures include mask-wearing ^11,12^, contact-free interactions ^13^, and restructuring of physical spaces ^14^. The use of masks, in particular, has been shown to be effective at reducing respiratory transmission of SARS-CoV-2, particularly when individuals in a potentially infectious interaction are wearing a mask ^15,16^. These population-wide measures still carry uncertainty since individuals are expected to behave uniformly irrespective of their disease status. As the scale of COVID-19 testing has increased, jurisdictions may also have an opportunity to consider implementing tactical mitigation strategies informed by testing.

Testing for infected status can, in theory, be used to initiate targeted isolation, identification and tracing of contacts, quarantining of contacts, and then selected testing of contacts ^17^. If done rapidly and at scale, this kind of targeted PCR-based testing can provide early detection of cases and help break new chains of transmission ^18,19^. The effectiveness of contact-tracing based control strategies hinges on the accurate identification and isolation of exposed and infectious cases. Slow return of test results (primarily) and false negatives (as a secondary factor) limit the effectiveness of test-based control policy ^20^. A complementary tactic is the strategic deployment of recovered individuals identified by serological tests in infection hot-spots to effectively dilute transmission events ^21,22^. Similar to the identification of infected individuals through PCR tests, immunity to SARS-CoV-2 is assessed through serological testing for protective antibodies ^23,24^. Population-wide interventions, testing efforts, the selective confinement or deployment of people contingent on their infection status, and inaccuracies and limited adherence to policies, combine to create a challenging landscape for the persistent control of outbreaks ^25^.

Non-pharmaceutical COVID-19 control until effective vaccines become available will necessarily involve periods of reduced social and economic activity; i.e., ‘business, but not as usual’. Control efforts are already generating hardship and could in the longer-term result in social unrest and increased mortality ^26,27,28^. Here we confront a joint problem: how to identify policies that aim to reduce fatalities arising from COVID-19 while also enabling economic engagement. Such a joint goal necessarily has a tension, can lead to dichotomous ‘extremal’ policies, and requires confrontation through careful modeling. We do so in two stages. First, we use optimal control to assess both health and economic outcomes in an SEIR disease model framework. There is a substantial and growing literature on optimal control for COVID-19, the bulk of which focuses on non-personalized release policies or policies that target age- or risk-stratified groups ^2,29,30,6.31,9^. Here, we identify optimal control policies to modulate interaction rates based on disease status identified by PCR and serological testing - unifying prior efforts centered on isolation and shield immunity. We find that intermediate policy outcomes can do nearly as well as strict public health scenarios, without incurring the severe costs as suppression-centered policies. However, optimal controls can be fragile, when applied in practice given that they rely on time- rather than state-based interventions; the consequence of mistiming interventions can be severe ^30^, Hence, guided by the optimal control analysis, we identify state-dependent policies similar to feedback control that provide actionable guidance for individual behavior. As we show, using population-wide PCR and serological testing as the basis for mitigation (rather than surveillance) has the potential to yield dual benefits if carried out at population scales: reducing COVID-19 transmission while enabling more individuals to return to work sooner and with fewer restrictions than would otherwise be possible.

## Results

### Optimal control framework for state-dependent contact rates policies that balance public health and socioeconomic costs

We develop an optimal control framework to identify policies that address the tension between decreasing contacts (that reduce new infections) with increasing contacts (that are linked to socio-economic benefits). We represent the epidemic using a Susceptible-Exposed-Infectious-Recovered (SEIR) nonlinear dynamic model (see Supplementary Information/Methods for complete details; see Figure 1). In doing so, the force of infection is influenced by state-specific contact rates *c_S_, c_E_, c_I_* and *c_R_* for susceptible, exposed, infectious and recovered individuals, respectively – these different levels form the basis for a control policy that directs individuals to interact at different levels depending on their test status.

**Figure 1:**
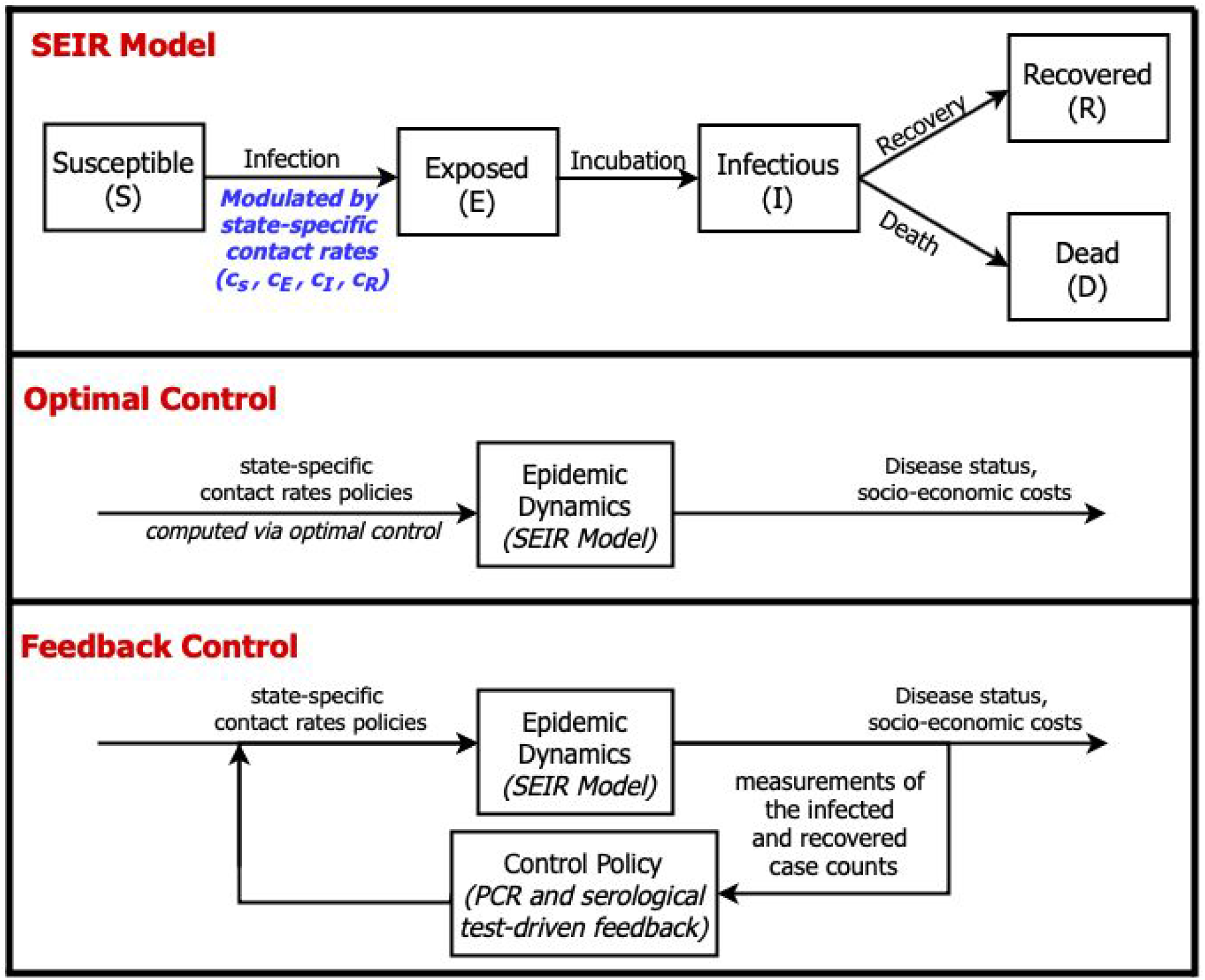
Epidemic dynamics with optimal and feedback control of disease-status driven contact rates. (Top) SEIR model schematic in which the force of infection is modulated by state-specific contact rates, see text and SIfor details. (Middle) Diagram of optimal control approach: contact rates are pre-specified given model structure and estimate of parameters and current conditions. (Bottom) Diagram of feedback control approach: contact rates are updated in real-time based on measurements of the infected and recovered case counts via testing surveillance.

In the optimal control framework, a set of state-specific contact rates are identified that minimize the appropriately weighted sum of what we term ‘public health’ and ‘socioeconomic’ costs. Public health costs are quantified both by average infected levels and cumulative deaths. Socioeconomic costs are quantified in terms of reductions in the total rate of interactions and by shifts in state-specific contact rates. The optimal control ‘solution’ is then a time-dependent set of disease-specific rates which are both shaped by and shape the epidemic itself (see SI/Methods for details on the gradient projection algorithm used to identify the solution). Note that we constrain the contact rate of exposed individuals to be equal to that of susceptible individuals given the challenges of timely identification of exposed individuals who are not yet infectious (and presumably have insufficient viral titer to be identified using screening tests; an issue we return to in the Discussion).

Figure 2 shows the results of comparing a baseline outbreak (i.e., neglecting public health costs, given weighting parameter ξ=0) to a full lockdown scenario (i.e., neglecting socioeconomic costs with 75% isolation for all, ξ>>1) and a balanced scenario with optimized contact-rates (i.e., corresponding to ξ = 1). As shown in Figure 2, in the baseline scenario, the disease spreads through the population leading to 94% cumulative infection (as expected given strength-size relationships for 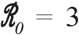). In contrast, a full lockdown scenario with 75% reduction in contact rates of all individuals after 60 days leads to a total outbreak size of 4% of the population. The optimal control solution in the balanced case ξ = 1 reveals a potential route to jointly address public health and socioeconomic cost. From the perspective of public health, the optimal control solution leads to 25% cumulative infections. In addition, the socioeconomic costs in the optimal control case are higher in the short-term but approach that of the baseline scenario in the long-term. Indeed, the effective reproduction number identified via an optimal control framework in the balanced scenario gradually reduces to sub-critical levels (close to an effective reproduction number, 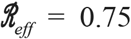) while gradually relaxing controls over time. The optimal control solutions are shown in Figure 2A. The optimal control solutions differ based on disease status, recovered individuals elevate their interactions, infectious individuals isolate, and susceptible individuals lock-down before gradually returning to pre-lockdown levels.

**Figure 2:**
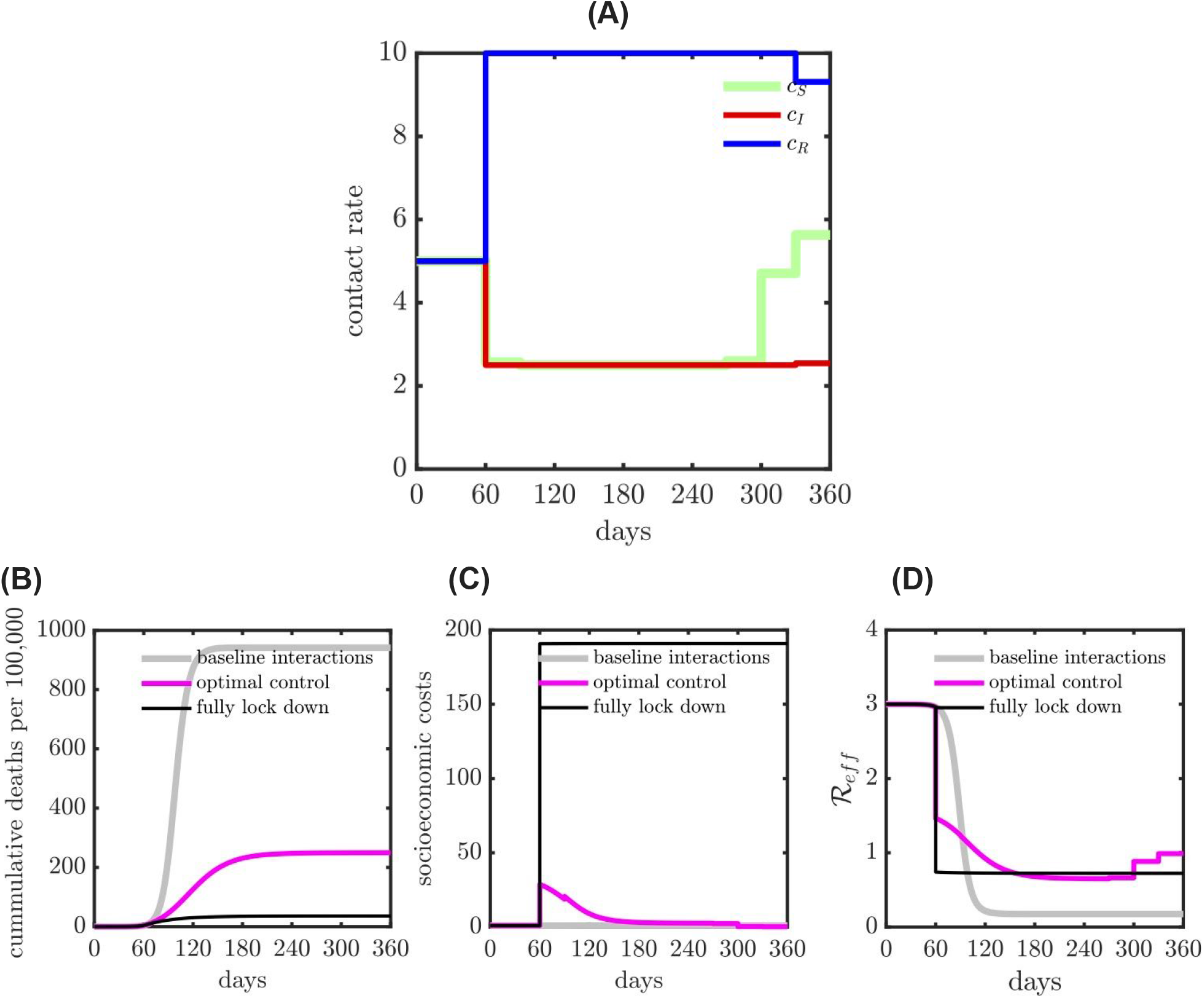
Comparison of health and economic outcomes of COVID-19 given various interventions: baseline interactions (i.e., no intervention); optimal contact rate intervention (balance both health and economic outcomes) and fully lock down intervention (applied to all the subpopulations) with 75% isolation efficiency. (A) The optimal contact rate with 50% isolation effectiveness and shield immunity level 2. (B) Cumulative deaths (health outcome) during the epidemic. (C) Socio-economic costs (economic outcome) during the epidemic. (D) Measure of effective reproduction number 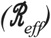 for different interventions during the epidemic.

### Personalized, test-based optimal control policies and their impact on public health and socioeconomic outcomes

In order to explore the mechanisms identified by the optimal control framework, we systematically modulate the effectiveness of isolation and evaluate its effect on the state-dependent optimal contact rates and disease dynamics. In practice, isolation effectiveness is influenced by availability, accuracy, and speed of testing as well as fundamental limitations on an individual’s ability to isolate (which can vary with socioeconomic and other factors). Figures 3A-C evaluate low, medium, and high efficiency of isolation spanning 25%, 50%, and 75% reduction in the contact rates of infected individuals, respectively. As is evident, the optimal control solutions for state-dependent contact rates vary significantly with isolation effectiveness; suggesting that COVID-19 response policies that can vary with disease status may open up new possibilities to balance public health and socioeconomic outcomes.

First, in the low (25%) or medium (50%) effectiveness cases, susceptible, exposed, and infectious individuals adopt the maximal level of isolation. Inefficient isolation of infectious individuals elevates risks of new transmission that are not outweighed by socioeconomic benefits. Notably, the optimal control solution includes an *elevated* level of interaction by recovered individuals. This finding recapitulates ‘shield immunity’ ^21,22^, insofar as recovered individuals are protected from re-infection over the course of the intervention. The elevated contacts of recovered individuals have multiple effects: both diluting interactions by susceptibles (and reducing transmission risk) and by increasing socioeconomic activity. In contrast, for sufficiently high levels of isolation efficiency (75%), the optimal control solutions suggest there is no need for a general lockdown. Instead, the combination of infected case isolation and shielding by the subpopulation of recovered individuals is sufficient to rapidly reduce and contain 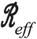 below 1, leading to a decreasing number of new infections. We note that irrespective of isolation effectiveness, balancing public health and economic outcomes drives below 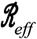 1, but not necessarily to 0 (albeit, given the constraints imposed by lockdown efficiency, such an extreme reduction may not even be possible), and eventually increased immunity permits an easing-off in restrictions yielding an increase in 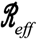 ^32^.

**Figure 3:**
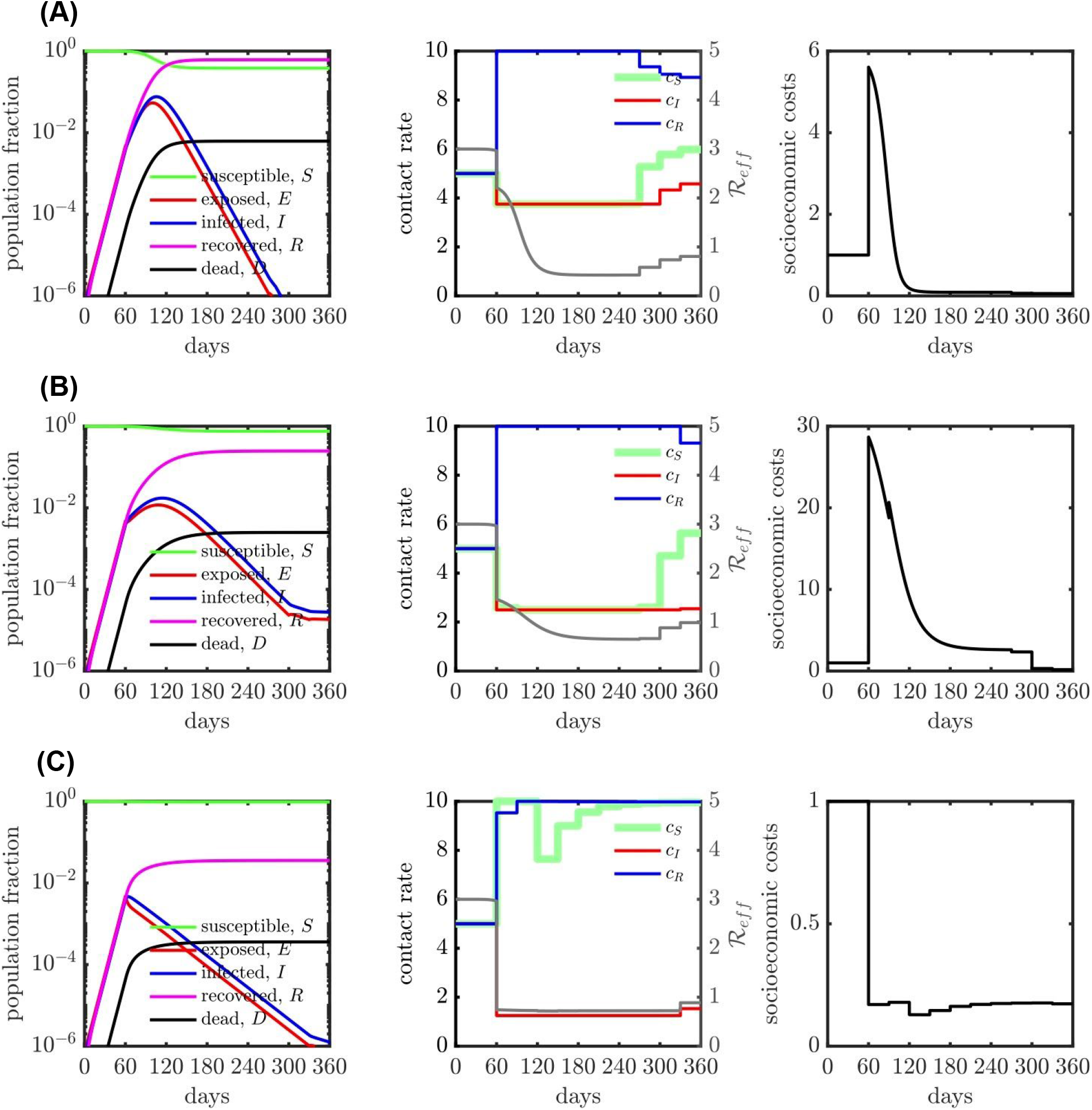
SEIR dynamics with contact rate interventions for various isolation efficiencies, (A) 25% isolation efficiency; (B) 50% isolation efficiency and (C) 75% isolation efficiency. The relative importance (ξ) is 1 for all the cases (A), (B) and (C). The contact rate interventions start at 60 days, people follow baseline (or normal) interactions before that. For all the isolation efficiency scenarios (three rows), the left panel shows the population dynamics given the optimal contact rate shown in the middle panel. The gray curve in the middle panel represents the measure of corresponding effective reproduction number 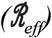. The right panel shows the corresponding socio-economic costs. See SI/Methods for additional scenarios.

### Sensitivity of optimal control approach to mis-timed implementation of policies

Despite its potential to balance public health and socioeconomic costs, a central drawback of optimal control solutions is the potential exponential growth of errors. Given the fact the COVID-19 dynamics are only partially observed (with significant uncertainty in the actual state), application of a policy requires an estimate of the time since epidemic initiation, what we term ‘epidemic age’. In order to evaluate the sensitivity of optimal control policies due to mis-timing, we first computed the optimal control policy for a system one month after an outbreak. However, instead of implementing the policy matched to the actual epidemic age, we enforce the optimal control policy 30 days later, i.e. at the end of 60 days after the start of the outbreak. Figure S7 shows the difference in the mis-timed control policy vs. the optimal control policy; as is evident the mistimed policy relaxes stringent lockdown when the optimal policy continues to lock-down. As a consequence the total deaths are far higher for 25% and 50% isolation efficiency (see Table 1). The mistimed policy, in effect, biases the system towards minimizing socioeconomic rather than public health costs. This significant difference in performance metrics demonstrates the potential shortcomings of implementing a policy based on optimal control. However we note that with a stringent isolation efficiency, delays are less problematic. The reason is that with efficient infected case isolation, both the mis-timed and optimal control policy could enable nearly all individuals to work, given that 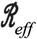 is held below 1 by infection isolation on its own.

Despite its fragility, we identify common features of the optimal control policy given variation in the effectiveness of infectious case isolation. First, the optimal control policy minimizes infected contact rates. The optimal control solutions also robustly identify an immune shielding strategy for recovered individuals, i.e., such that recovered individuals elevate their interactions to the maximum possible relative to baseline. Importantly, differences in the optimal control policy are primarily centered on identifying a switch point in contact rate level for the susceptible population. From Figures S4, S5 and S6, we observe the switch point as a function of time, showing that irrespective of isolated case effectiveness and shield immunity constraints, the increase in susceptible contact rates happens later in the lockdown period. Switchover points correspond to times when the infection prevalence is relatively low compared to the recovered population. This observation provides the basis for a feedback, rather than optimal, control policy.

**Table 1:**
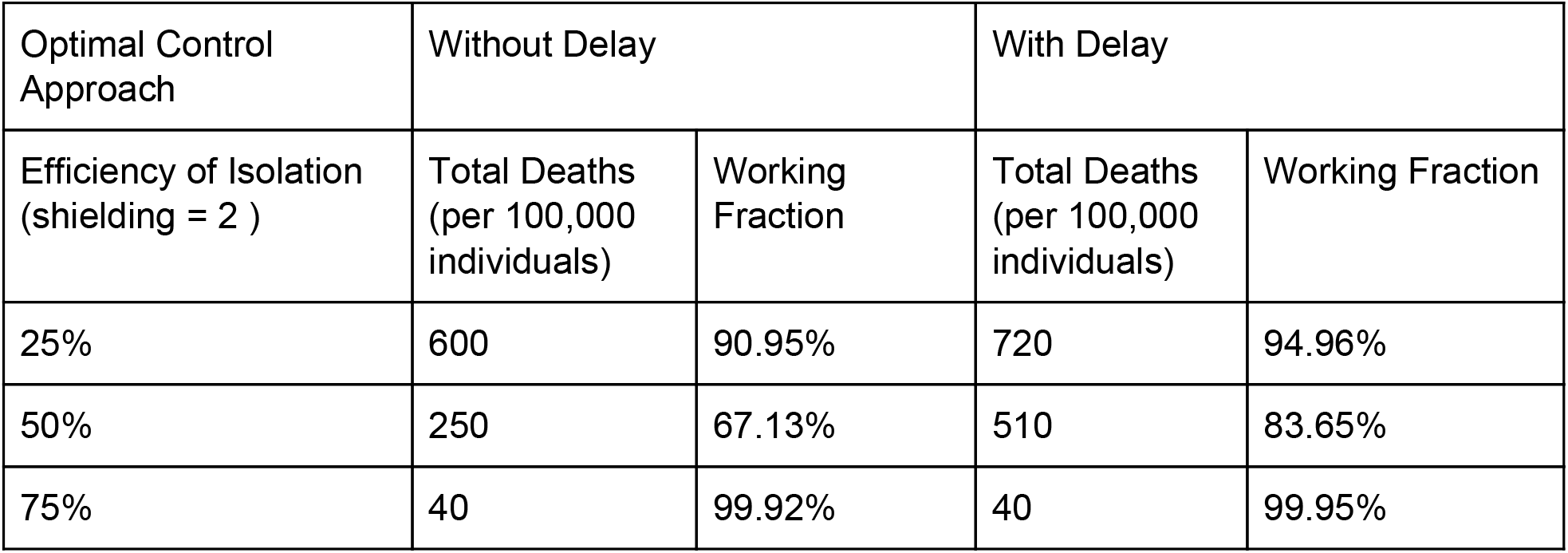
Comparison between optimal control approach for contact policy with and without delay. The comparisons are made for isolation efficiencies of 25%, 50% and 75%, with performance metrics of total deaths (per 100,000 individuals) and working fraction. The total death is significantly higher for the system with delay when the isolation efficiency is not 75%, suggesting poor robustness to delay.

### Feedback-control policy for balancing public health and socioeconomic costs

We propose the use of a feedback control policy adapted from emergent features of the optimal control policy solutions: (i) infectious individuals isolate as far as is possible; (ii) recovered individuals increase their activities as much as possible, i.e., akin to shield immunity. Hence, we set out to identify a system-dependent change in the contact rate of susceptible individuals, separating lockdowns vs. return-to-work. In practice, we identify a critical curve in *I-R* plane (i.e., infected-recovered cases plane) via a genetic algorithm, such that the recommended behavior of susceptible individuals is dictated by surveillance-based estimates of infectious and recovered individuals (see Supplementary Information for more details).

Figure 4 summarizes the results of the feedback control policy. From a policy perspective, for both 25% and 50% isolation effectiveness the feedback control policy identifies a switch between lock-down and return-to-work when there are significantly more recovered individuals than infectious individuals (approximately 100x as many). Hence, circulating case levels should be relatively low relative to recovered individuals before shifting from lockdowns to re-openings. We also examined the same scenarios while assuming recovered individuals return to work but without the beneficial effects of shield immunity. As is apparent, the policies are more stringent, with the critical relationship shifting to requiring that there are more than 350 times as many recovered individuals as infected individuals.

Critically, the performance of the test-driven feedback policy is nearly identical for the performance metrics with or without mis-timing (see Table 2). This finding implies that state-based approaches will be less likely to have exponentially mis-timed applications, and reinforces the need for population-scale testing for both active infections (via PCR) and prior infections (via serological and/or antigenic based testing that evaluates protective immune responses). We note that a simple policy with only two states - ‘lockdown’ and ‘open’, respectively corresponding to minimum and baseline contact rates for the susceptible cases, would be easier to implement than one with continuous ‘phases’ or state changes. In Figure S9, we document the generalizability of results given variation in infected case isolation and the level of shield immunity.

**Table 2:** Comparison between feedback control approach for contact policy with and without delay. The comparisons are made for isolation efficiencies of 25%, 50% and 75%, with performance metrics of total deaths and working fraction. Both performance metrics are nearly identical for all efficiencies, suggesting no significant effect of delay on the system.

| Feedback Control Approach | Without Delay |  | With Delay |  |
| --- | --- | --- | --- | --- |
| Efficiency of Isolation (shielding = 2 ) | Total Deaths (per 100,000 individuals) | Working Fraction | Total Deaths (per 100,000 individuals) | Working Fraction |
| 25% | 620 | 89.98% | 620 | 89.94% |
| 50% | 250 | 62.40% | 250 | 62.12% |
| 75% | 30 | 99.90% | 30 | 99.90% |

**Figure 4:**
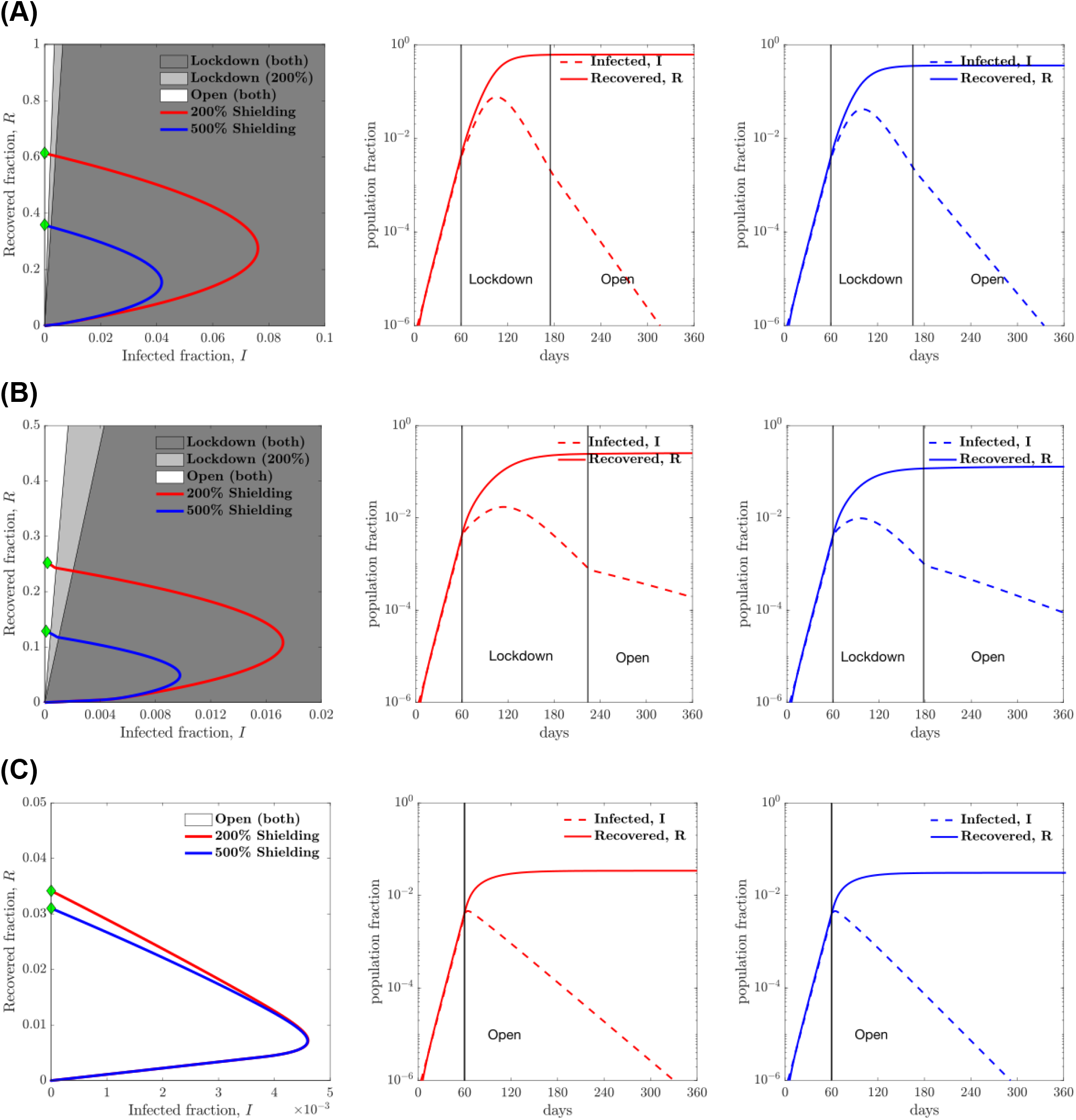
Heuristic state feedback intervention policies varying with isolation efficiency: (A) 25% isolation efficiency; (B) 50% isolation efficiency and (C) 75% isolation efficiency. The shielding levels considered here are 2 and 5 times the base contact rate (i.e., 200% shielding and 500% shielding respectively). An optimal line divides the plane into two regions which determines the optimal contact rate for the susceptible population for the current infected and recovered cases. The optimal policy in the dark grey region is lockdown for both shielding levels while the system with a higher level of shielding (level of 5) is open in the light grey region. The phase plots (red and blue curves) show the evolution of the infected and recovered case fractions over the period of 360 days, while applying the control strategy described above for shielding levels of 2 and 5 respectively. See SI/Methods for a larger set of plots with increments of 5% in isolation efficiency and for shielding levels of 2, 3, 4 and 5. The second and third plots show the infected and recovered cases for shielding levels of 2 and 5 respectively as functions of time. The vertical lines mark the time instances at which the policy is implemented with lockdown imposed and also the time at which it is lifted. For the case of isolation efficiency of 75%, no lockdown is needed at all for the susceptible population.

### Concluding remarks

We have developed a linked series of optimal- and feedback-control analyses to evaluate the effectiveness (and benefits) of modifying contact rates for managing the COVID19 pandemic from both health and economic perspectives. Throughout, our central goal was to optimize the interactions between individuals (in different compartments) so as to achieve a defined balance between public health and economic outcomes. By explicitly incorporating contact rates as the control variables in a SEIR model, we were able to identify optimal control policies that could, in theory, significantly reduce expected infections (and fatalities) while reducing the negative socioeconomic costs of sustained lockdowns. Optimal control policies are unlikely to be applied in practice, given the potential exponential mis-specification of policies over time. Hence, we leveraged insights from the optimal control solutions to guide a feedback control approach that performs nearly as well as the optimal control approach with significant improvements in robustness given uncertainty in estimating the epidemic state. Collectively, our control policies indicate that infected individuals should be isolated (as effectively as possible), recovered individuals should be encouraged to return to work (given benefits accrued via shield immunity), while the release of other individuals from lock-down should be guided by the epidemic state. The transition from lock-down to return-to-work occurs when circulating case loads are far lower than recovered case counts; with the scope of the epidemic sharply controlled by infection case isolation. A combination of policies, e.g., mask-wearing, physical distancing, will help to reduce transmission risk for individuals who do return to work.

We recognize that the SEIR framework is intentionally simplified. The epidemic model does not account for process or observational noise, analytic test features, heterogeneity, stratified risk, asymptomatic cases, and detailed elaboration of severe cases. By reducing the model complexity we have tried to shed light on the general problem of balancing public health with socioeconomic outcomes. In doing so, we have highlighted a middle ground between dichotomous outcomes that focus on public health or socioeconomic costs solely. Central to our work is the presumption that testing is sufficient to estimate both the circulating case counts and the number of recovered individuals. In doing so, serological testing is key. Given under-testing, the calibration of reported cases requires either model-based inference or serological test comparisons to estimate the ascertainment bias ^33^. Collectively, we remain closer to the beginning than the end of the Covid-19 pandemic. As we have shown, accelerating the slow-down of transmission while restoring economic activity may be enabled by both personalized, test-driven, policies.

### Data availability

All simulation and figure codes used in the development of this manuscript are available at https://github.com/WeitzGroup/COVID_Control_2020

## Data Availability

All simulation and codes used in the development of this manuscript are available at https://github.com/WeitzGroup/COVID_Control_2020

https://github.com/WeitzGroup/COVID_Control_2020

## Acknowledgements

We thank Weitz Group team members for comments and feedback. This work was supported by grants from the Army Research Office (W911NF1910384), National Institutes of Health (1R01AI46592-01), and the National Science Foundation (1806606 and 2032082).

## Supplementary Information

### I. METHODS

#### A. Model and parameters

The epidemic model analyzed here is an ’SEIR’ model, including SEIR dynamics [1], susceptibles *S*, exposed *E*, infectious *I*, recovered *R*, with the total number of fatalities denoted by *D*. The *force of infection* is the contact rate of infectious individuals multiplied by the probability that the interaction is with a susceptible person multiplied by the probability that the event leads to an infection. Let *c_S_*, *c_E_*, *c_I_* and *c_R_* equal the contact rate of *S, E, I*, and *R* individuals respectively. The force of infection is

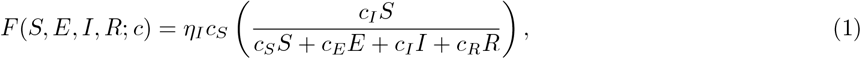

where *η_I_* is the measure of disease transmission effectiveness from *I* class to *S* class[2]. Suppose all the contact rates are equal to a baseline contact rate *c_B_*. Assuming relatively few fatalities per-capita, then the total population is approximately constant at *N*, such that the force of infection can be approximated as *F* ≈ *η_I_c_B_*(*I*/*N*), which recovers the standard SEIR model by defining *β* = *ηc_B_* as *infection rate*. In the generalized case, the model is given by the following system of nonlinear differential equations

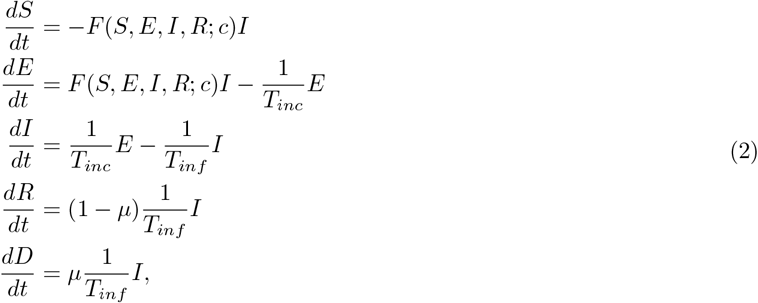

where *F(S,E,I, R; c)* is the force of infection given in Eq. 1, and *T_inf_* is the infectious period, *T_inc_* is the incubation period, *μ* is the case fatality ratio for infected individuals, see SEIR model schematic in Fig. S1.

**FIG. S1:**
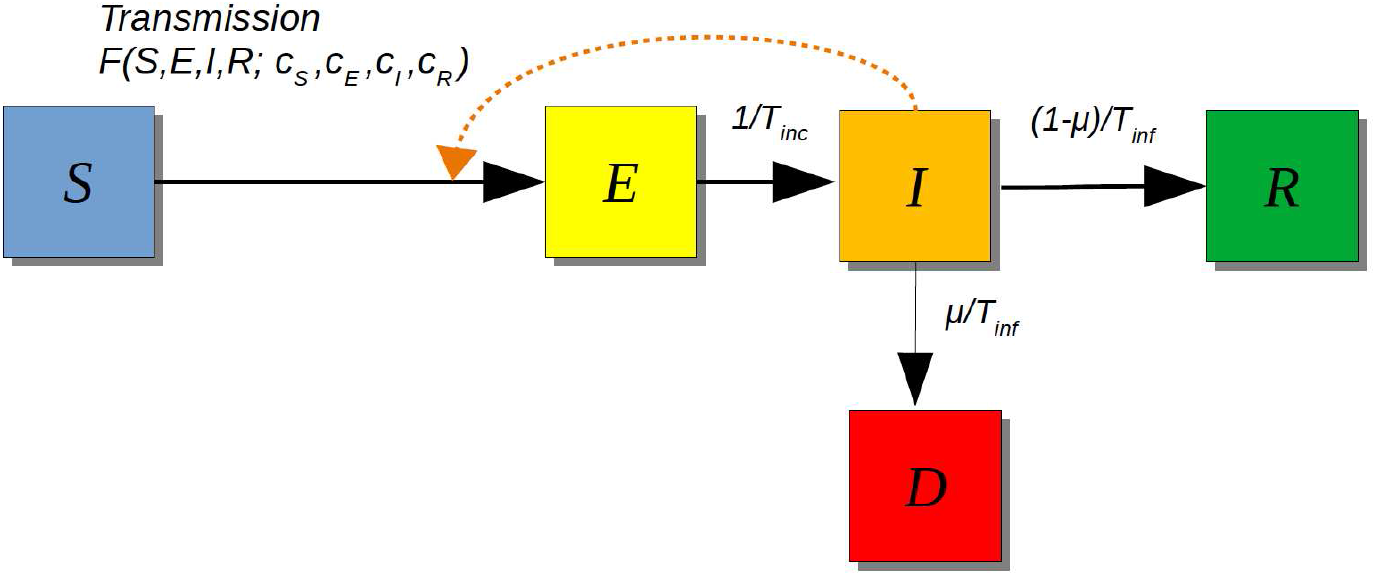
SEIR Model Schematic. The interactions between susceptible and infected individuals lead to newly exposed cases. These interactions are modeled by the force of infection *F*(*S, E, I, R; c*). The exposed individuals undergo an incubation period (*T_inc_*) before the onset of infectiousness. Infectious individuals will either recover (and develop protective immunity) or die after an infectious period (*T_inf_*), see model equations in Eq. 2.

At the start of an outbreak (*t* = 0), the contact rate of each individual is *c_B_*. The basic reproduction number 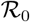 of this compartmental model is

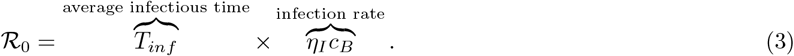

The assumed model parameters used in the model are shown in table S3.

##### Initial conditions and contact rate

The model assumes an initial total population of 1, 000, 000 (one million) denoted as *N*_0_. An initial outbreak is seeded in this population given one infected individual. The simulation is run for two months (60 days) with fixed baseline contact rate (all *c*’s are equal to *c_B_*) - we use this time point (which we denote time *t*_0_ in our simulations) as the time at which intervention policies might be applied. The individual behaviors (or activities) are quantified by contact rates, *c_S_*(*t*), *c_E_*(*t*), *c_I_*(*t*) and *c_R_*(*t*), thus the control of contact rates can be mapped to intervention strategies. For examples, when cs goes down that means shelter-in-place, when *c_E_* or *c_I_* goes down that can mean quarantine and isolation. When there is no intervention during the epidemic, the population dynamics of SEIR model is shown in Fig. S2.

**FIG. S2:**
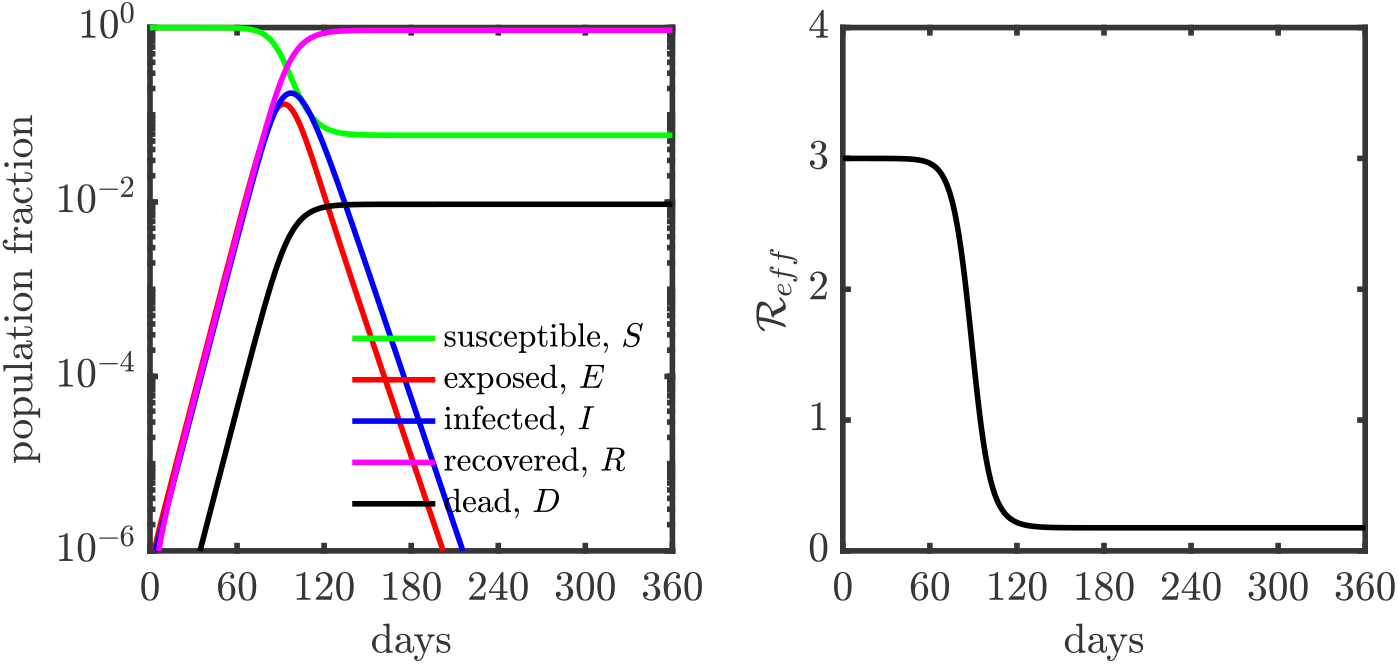
Population dynamics of SEIR model without control, *i.e*., all *c′s* are equal *c_B_* from to *t*_0_ *t_f_*. (Left) Population dynamics of SEIR model without control. (Right) Epidemic growth as measured by the effective reproduction number (*R_eff_*), the basic reproduction number 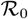 is 3.

We denote *c*(*t*) = (*c_S_*(*t*)*,c_E_*(*t*),*c_I_*(*t*),*c_R_*(*t*))*^T^* as the contact rate vector at time *t*, where (·)*^T^* denotes a matrix transpose. The effective reproduction number 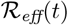 is

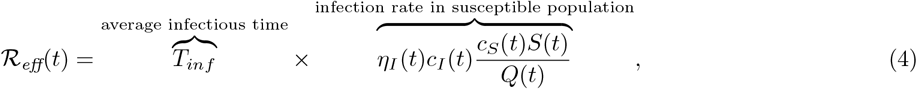

where *Q*(*t*) = *c_S_*(*t*)*S*(*t*) + *c_E_*(*t*)*E*(*t*) + *c_I_*(*t*)*I*(*t*) + *c_R_*(*t*)*R*(*t*).

#### B. Optimal control formulation

Here, we aim to optimally deploy public health control strategies (via control of the contact rates) that minimizes deaths during the outbreak while keeping the socioeconomic costs low. The baseline average of contacts at time *t*, *Q*(*t*), equals *c_B_N*(*t*), where *c_B_* is the baseline contact rate and *N*(*t*) = *S*(*t*) + *E*(*t*) + *I*(*t*) + *R*(*t*) is the total number of alive individuals The socioeconomic costs may result from two consequences of contact rate interventions: (1) loss of essential connections; (2) shifting of roles relative to baseline, as quantified by variation in contact rates by testing status. Given the intervention policy *c*(*t*) at time *t*, we quantify the socioeconomic costs as 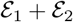, where 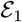 and 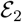 may take form of

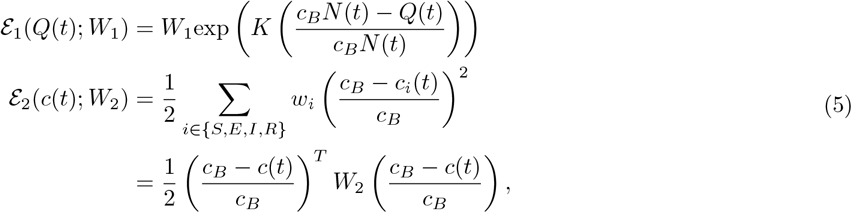

where *W*_1_ ≥ 0, *W*_2_ = diag(*w_S_,w_E_,w_I_,w_R_*) ≻ 0 are weight parameters, and *K* is a constant shape parameter for the exponential function. In the absence of any strategy, there are few deaths. This would mean that *N*(*t*) ≈ *N*_0_. For the sake of simplicity, 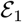 is approximated as *W*_1_exp (*K* (*c_B_N*_0_ − *Q*(*t*)) /*c_B_N*_0_). For every *t*, we restrict the contact rate vector *c*(*t*) ∈ [*c_min_, c_max_*]^4^, where *c_min_* and *c_max_* are the minimum contact rate and maximum contact rate. The set *C* = [*c_min_, c_max_*]^4^ is a convex and compact set in ℝ^4^. We define the space of *admissible controls*, denoted by A, as the set of Lebesgue-measurable functions *c*: [*t*_0_*,t_f_*] → *C*. The (dimensionless) cost functional is written in the following Bolza form,

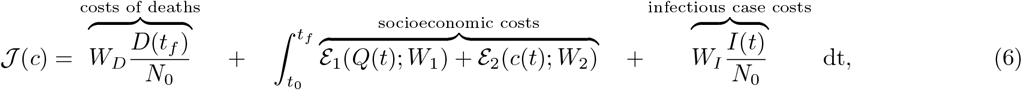

where *W_I_* ≥ 0 and *W_D_* ≥ 0 are weight regulators. The optimal control problem is

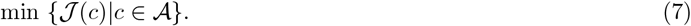

The weight parameters used in Eq. 6 are shown in table S4, these weight parameters are set to balance the scale of different cost components in Eq. 6.

However, frequent and continuous updates bring in practical issues in terms of (1) establishing how subtle changes in contact rates actually translate into applicable changes in individual behaviors (*i.e*., a continuous strategy can not be practically implemented), and (2) individual fatigue (and reduced adherence to proposed measures) due to frequent behavioral adjustments (*i.e*., frequent update is also impractical). To address these concerns, we need to modify the space of controls 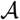.

To begin with, we fix the full time horizon of control as [*t*_0_*,t_f_*]. For example, *t*_0_ is fixed to 60 days after outbreak (see Sect. I A) and *t_f_* to 360 days after outbreak, where we set the timing of the outbreak to 0. We only consider an intervention policy which is updated a finite number of times in the interval *t*_0_*,t_f_* ]. These epochs are denoted by *t*_0_ *<t*_1_ *<t*_2_ *<* … < *t_M_* = *t_f_*. Let 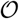 denote the corresponding partition of [*t*_0_, *t_f_* ], and denote by 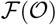 the space of functions *c*: [*t*_0_*,t_f_*] → ℝ^4^ which have constant values in the intervals [*t_m_*_−1_*,t_m_*), *m* = 1, *2,…*. The resulting optimal control problem is

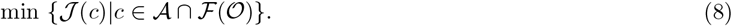

In most cases, the exposed individuals are unaware of their infection status, so it’s impractical to control their behaviors separately from the susceptible individuals. Hence, we view them as susceptibles for the purpose of policy setting and set *c_E_*(*t*) = *c_S_*(*t*) ∀*t* ∈ [*t*_0_,*t_f_*]. In doing so, the contact rate vector c(*t*) actually is a three dimensional vector consisting of *c_S_*(*t*), *c_I_*(*t*) and *c_R_*(*t*).

#### C. Optimization algorithm

We follow Polak [3] who developed a general framework for computational optimal control based on optimization on Hilbert spaces. We first discuss the conceptual algorithm in an abstract setting of infinite-dimensional spaces, and then provide approximation details in the sequel.

Let 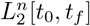 denote the space of equivalence classes of square integrable functions from [*t*_0_*,t_f_*] into ℝ^n^ (*n* = 3 in our case). We use the notation ||·|| and 〈·, ·〉 for the norm and scalar product in Euclidean space (e.g. ℝ^n^), and use 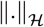 and 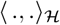 for the norm and scalar product in a Hilbert space 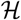. For example, given *u*, 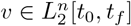 (a Hilbert space), the norm ||·||_2_ and scalar product 〈., .〉_2_ are: 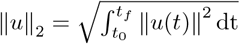; 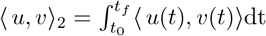.

In the optimal control problem under consideration, we define the state variable of the system, *x* ∈ ℝ^5^, by *x* = (*S,E,I,R,D*)*^T^*. We also define the control variable, *c* ∈ ℝ^3^, by *c* = (*c_S_,c_I_*,*c_R_*)*^T^*. The dynamic equation of the system, Eq. 2, can be rewritten in a compact form:

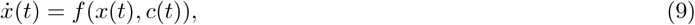

where dot represents differentiation with respect to time *t*, and *f*(*x*(*t*)*, c*(*t*)) is the right hand side (RHS) of equations 2. The cost functional defined by Eq. 6 can be written succintly as

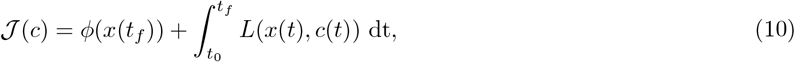

where 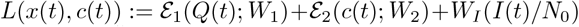 and *ϕ*(*x*(*t_f_*)) = *W_D_*(*D*(*t_f_*)/*N*_0_). The costate (adjoint) variable, λ(*t*) ∈ ℝ^5^, is defined by the following equation,

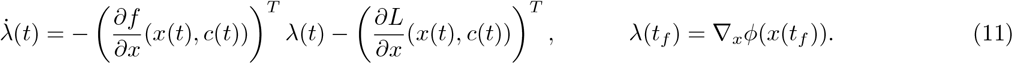

Next, recall the optimal control problem defined by Eq. 8, where the cost functional (performance integral) 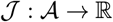 is defined by (10). We will propose a gradient-descent algorithm for solving this problem, where the gradient 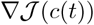 is given by the following equation,

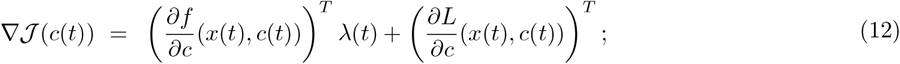

see [3–5].

The principal elements of most gradient-descent algorithms (including the one described here) are the descent direction and the stepsize. For our problem, the descent direction is based on 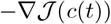; in fact, it is the projection of 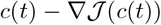 into the constraint-set *C* = [*c_min_,c_max_*]^3^ as defined in Sect. IB. The stepsize that we use is the Armijo stepsize (see [3]) described in the sequel, which gives an approximate line minimization of 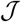 in the computed descent direction from the control *c*(*t*).

Consider first the conceptual algorithm. In its formal presentation by Algorithm 1, below, we use the term “compute” in a conceptual sense, since the “computations” refer to elements in the Hilbert space 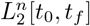. Also, we use the notation 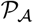 to mean the projection from 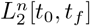 into the space 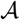.

##### Algorithm 1: Conceptual

Given a control iteration 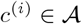, compute from it the next iteration, 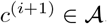, as follows.

*Parameters:* Constants *α* ∈ (0,1) and *β* ∈ (0,1). (*These parameters play a role in the computation of the step size.)*

1. Compute the state trajectory *x*(*t*) and costate trajectory λ(*t*) by Eqs. 9 and 11, respectively.
2. Compute 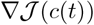 by Eq. 12.
3. (*Here we define, for all* 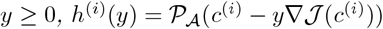). Compute *ℓ*(*i*) > 0 defined as follows,

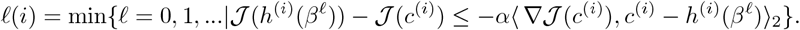
4. Set γ(*i*) = *β*^ℓ(^*^i^*^)^, and set

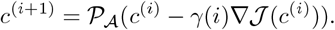

We remark that 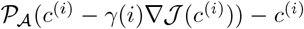 is descent direction for 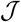 from *c*^(^*^i^*^)^, and γ(*i*), the Armijo stepsize, provides an approximate-line minimization in that direction.

The main modification of Algorithm 1 towards implementation is in numerical solutions of the state equation and costate equation. Furthermore, the time-interval [*t*_0_,*t_f_*] has to be discretized by a suitable grid in order to adequately represent various time-dependent functions such as *c*^(^*^i^*^)^(*t*) and 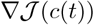. All of this can be achieved by a common grid, 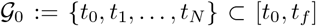. Furthermore, the projection 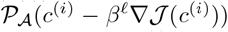 in Step 3 of Algorithm 1 can be computed as follows: For a given grid-point 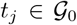, project the point 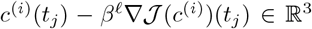 into the set *C*, where *C* = [*c_min_,c_max_*]^3^ ⊂ ℝ^3^. This can be done by a simple co-ordinate projection since *C* is a box. Denote the result of this pointwise projection by 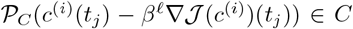, and observe that the function comprised of a zero-order or first-order interpolation of these points can serve to approximate the functional projection 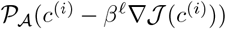.

These modifications of the conceptual algorithm give an implementable version for it. The resulting implementable algorithm falls in Polak’s framework of numerical optimal control [3] which includes results pertaining to asymptotic convergence.

As noted, the control c(*t*) must be piecewise constant and maintain constant values for substantial periods. At the same time, the state trajectory typically is continuous and not piecewise constant. Therefore, it is natural to maintain the present framework of continuous-time optimal control and not to consider the problem in the setting of discrete-time systems. To do that, we have to consider a form of projection of the control functions, c(*t*), into the space of piecewise-constant functions. We next describe the specific way we did that in our simulations.

Consider an increasing set of points {*t*_0_ = *t*_0_ < *t*_1_ < *t*_2_ < … < *t_M_* = *t_f_*}, and the corresponding partition of the time-interval [*t*_0_, *t_f_*) by the subintervals [*t_m_*_−1_, *t_m_*), *m* = 1,…, *M*. Denote the interval [t_m-1_, *t_m_*), *m* = 1,…, *M*, by Δ*_m_*; right-close the last interval to be Δ*_M_* = [*t_M_*_−1_, *t_M_*], and denote the partition by 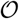. Let *ψ_m_*(*t*), *m* = 1,.. ., *M* denote the indicator function of the subinterval *ψ_m_*, namely *ψ_m_*(*t*) = 1 for *t* ∈ *ψ_m_* and *ψ_m_*(*t*) = 0 otherwise. A continuous-time signal can be approximated over an interval by a constant equal to its average value over that interval. Given a continuous-time policy *c*: [*t*_0_,*t_f_*] → ℝ^3^, the piecewise constant reconstruction is

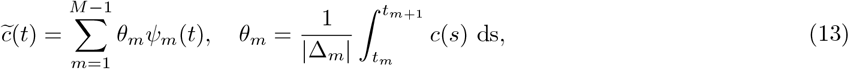

here *θ_m_* ∈ ℝ^3^. Let 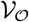 denotes the operator comprising this piecewise- constant reconstruction procedure. Then Eq. 13 can be written as 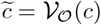. Now we modify the algorithm by replacing the projection operator 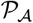 in Step 3 by the operator 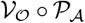, comprised of 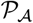 followed by 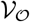.

In summary, the implementable algorithm that we use consists of the following modification of Algorithm 1:

1. Compute *x*(*t*) and λ(*t*) by numerical integrations using the forward-Euler method.
2. Compute 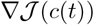 only at the points *t_j_* on the grid 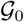.
3. Redefine *h*^(^*^i^*^)^(*y*) as 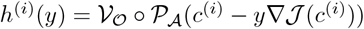.
4. Replace 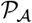 by 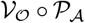.

A flow chart of the algorithm is depicted in Fig. S3.

##### Simulation details

The model assumes an initial total population of 1, 000, 000 (one million) denoted as *N*_0_. An initial outbreak is seeded in this population given one infected individual. The simulation is run for two months (60 days) with fixed baseline contact rate (all *c*’s are equal to *c_B_*) - we use this time point (which we denote time *t*_0_ in our simulations) as the time at which intervention policies might be applied: we have *c*(*t*) = *c_B_* ∀*t* ∈ [0, *t*_0_], and we set the initial state to *x*_0_ = *x*(*t*_0_).

For the algorithm, we set *α* = 0.1 and *β* = 0.5, the grid’s time increments to 0.05, Δ*_m_* = 30 days, *t*_0_ = 60 and *t_f_* = 360 days. For the uncontrolled part of the simulation we take *c*(*t*) = *c_B_* for all *t* ∈ [0, *t*_0_]. The term 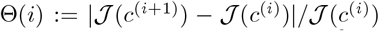 acts as a convergence indicator for stopping the algorithm. The algorithm is terminated whenever ϴ(*i*) ≤ 10^−6^.

#### D. Optimised time-dependent contact rate

One way of estimating the economic impact is measuring the *fraction of working-days recovered, i.e*., how many days are people working. We assume that if a person has a contact rate less than the base contact rate (*c_B_*), it implies a proportional reduction of in-person contacts that have a direct economic benefit. For example, for a susceptible individual, the working fraction at day *t* can be computed as 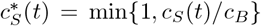, where *c_B_* is the baseline contacting rate. This ensures that contact rates above the baseline does not correspond to an increase in hours worked. For a given contacting rate policy *c* = (*c_S_*, *c_E_*, *c_I_*, *c_R_*)*^T^*, the working days restored in the period [*t*_0_, *t_f_*] is

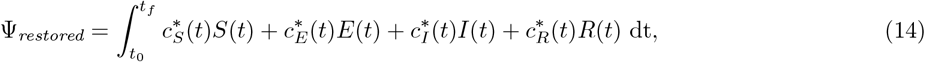

where 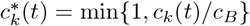, *k* ∈ *{S,E,I,R}*, is the working fraction of an individual in subpopulation class *k* at day *t*. When there is no policy intervention, the contacting rate of each individual is *c_B_* for all time, the working days will be maximized, which can be written as

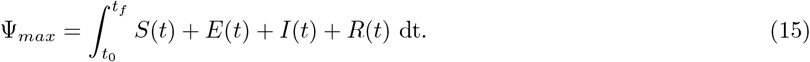

**FIG. S3:**
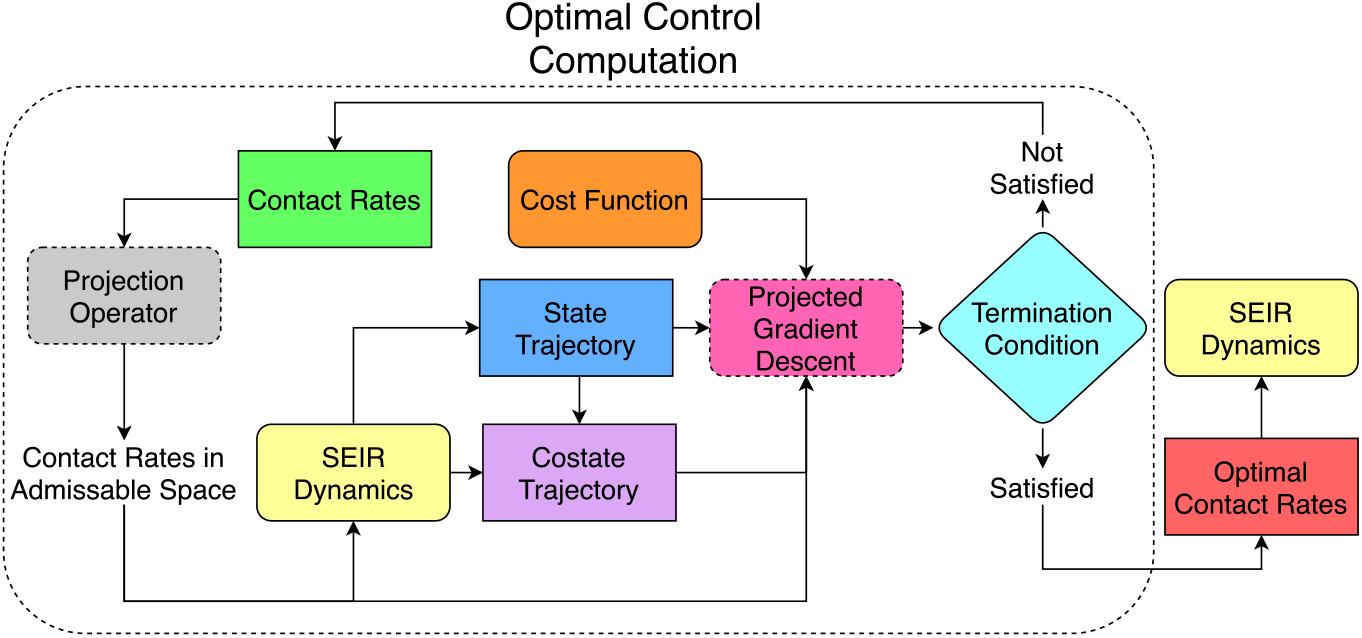
Flowchart of optimal contact rate computation. The contact rates are projected into the allowed subspace, and then fed to the system dynamics to generate the state and costate trajectory. Using the cost function provided and the state and costate trajectories, a projected gradient descent algorithm is employed to update the contact rates. If the termination condition is satisfied, these new contact rates are accepted as the optimal contact rates. Otherwise, the contact rates are replaced with the computed values and the loop is rerun.

Then, the fraction of working days restored can be written as

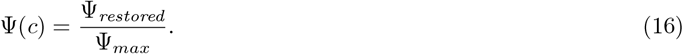

Economic costs are proportional to 1 − Ψ(*c*); such that economic impacts of policies are minimized as Ψ(*c*) approaches 1

##### Varying the relative importance of the illness-related cost vs. socioeconomic costs

The relative importance given to the socioeconomic impact of the epidemic and the cost associated with deaths and infection spread is critical to design intervention policies. From Eq. 6, we note that the cost function 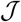 can be decomposed into the two parts, socioeconomic costs 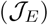 and costs of infectious diseases 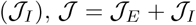, where

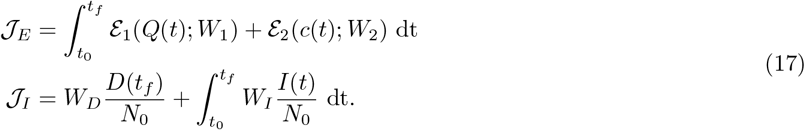

In order to explore how to manage and control disease outbreak with low socioeconomic costs, we need to balance the importance of these two seemingly conflicting objectives. To this end, we define a new cost function where the relative importance of 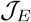 and 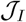 can be parameterized. With this parameter, optimal policy decisions can be made based on the relative importance assigned to the socioeconomic effect and public health impacts. Define the relative weight ratio as ξ and the altered cost function is 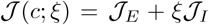. As ξ increases, the relative importance of costs associated with death and spread of infection are increased vis-a-vis the socioeconomic impact and vice versa. The optimal intervention policies with different relative importance ratios (ξ = 10^−4^, 1, and 10^4^) vary with isolation efficiencies (25%, 50%, 75%) are shown in Figs. S4, S5, S6.

For example of balance case (ξ = 1) in the case of 50% isolation efficiency, Fig. S5 (middle) shows that the way of managing the epidemic while considering the socioeconomic impact is to isolate all the infected cases and enhance the interactions of recovered cases (*i.e., shielding*, [6]), the susceptible cases are stay home first and slowly go back to normal towards the end of intervention period. Using the optimized intervention policy (*i.e*., optimal contact rate), the reduction in final deaths (*D*(*t_f_*)) is substantial (~ 75%). The fraction of working-days recovered is about ~ 66%. The results suggest advantages of isolation of infected cases and shielding enhancement of recovered cases.

#### E. Inferring state-dependent heuristic policies from optimal control solutions

In the previous sections, an optimal policy based on contact rates of different sub-populations was derived. However, as is often the case with optimal control solutions, these policies are open-loop, that is, once computed, they are prescribed to the system without feedback. Therefore, open-loop policies can have large sensitivities to modeling errors and signal-delays in the loop, especially if the system is unstable, as is the case with dynamic models of epidemics near the disease-free equilibrium at the onset of an outbreak. To address this, we design a feedback law for optimizing a ‘reasonable’ performance metric which, though different from the one of the optimal control problem, gives similar respective measures of two important performance metrics: total mortality (*D*(*t_f_*)), and working fractions (Ψ). In Sect. IF we verify this point by comparative simulations of the optimal control solution and the feedback control system, and not surprisingly, the sensitivity of the former with respect to a one-cycle (30-days) delay is much larger than that of the latter.

The optimal control solutions reveal a general structure that guides us in the choice of the feedback control law. The patterns observed in the computed optimal-policy solutions (see Figs. S4, S5, S6) suggest that the contact rates for the infected cases should be the smallest possible while the recovered population should be used as shields. Further, they display switch overs between the low and high contact rates for the susceptible populations, and these tend to occur towards the end of the lock-down period in a way that depends on the numbers of infected and recovered individuals.

To find the state feedback policy, the *I-R* (Infected-Recovered) phase plane is divided into 2 parts - one where the susceptibles need to isolate and another where they can get back to work. If the number of infected cases is high, the susceptible population should isolate to reduce the infection spread. For simplicity, the *I-R* plane is divided by a line, though better classification boundaries may exist. On one side of the line, the contact rate for the susceptible population is set to the minimum while on the other side, it is equal to the baseline (corresponding to opening the system).

The optimal slope of the dividing line is found via a *genetic algorithm* [7] using `matlab’s` built-in optimization function `ga`, with the maximum generation number (set to 30) serving as the stopping criterion [8]. The intercept of the line is fixed to a small negative number (10^−3^) as the results are not affected by the intercept. The slope parameter is called *θ*, and the following cost function is minimized:

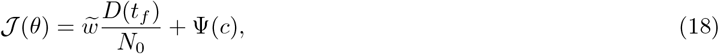

where *D*(*t_f_*)/*N*_0_ gives the normalized casualties, Ψ(*c*) is the fraction of working-days restored and 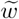 is a weight to bring the two terms on the same scale (see table S4). The output of the genetic algorithm is *θ*^*^, the slope of the optimal dividing line. We assume that the divided plane on the side of the line with the origin, *i.e*., zero infected and recovered cases, is the side where the susceptibles are free to work, while the divided plane on the other side corresponds to lockdown. The simulation is run for different isolation and shielding levels and the compiled results are shown in Fig. S9. The isolation efficiencies vary from 25% to 75% while the shielding ratio varies from 2 to 5.

We note that as the efficiency of isolation increases, the susceptible population can be let out for a longer duration towards the end of the epidemic. This is because the reduced contact rate of the infected population reduces the spread of infection. If the isolation efficiency is 65% or more, there is no need for the susceptible population to be in lockdown. Similar observations can be made for the shielding ratio. While the effects of it are not as dramatic, an increase in shielding ratio decreases the force of infection, resulting in a a lower time necessary for lockdown.

#### F. Sensitivity of optimal control and feedback control to mis-timed implementation of policies

We compute an optimal control for a system with a delayed input. We do not assume any modeling uncertainty, and attribute the perturbation only to the delayed application of the computed control. Suppose that an optimal control is computed for a scenario where its application is slated to commence 30 days after the outbreak of an epidemic, but the computed input control is uniformly delayed by 30 days. Then, by simulating the epidemic with a delayed policy, we can compare total deaths and working fraction for the delayed system with the results obtained without delay and with the same initial conditions.

**TABLE S1:** Comparison between optimal control approach for contact policy with and without delay. The comparisons are made for isolation efficiencies of 25%, 50% and 75%, with performance metrics of total deaths and working fraction. The total death is significantly higher for the system with delay when the isolation efficiency is 25% and 50%, suggesting poor robustness of the computed optimal control to input-delay.

| Optimal Control Approach | Without Delay |  | With Delay |  |
| --- | --- | --- | --- | --- |
| Efficiency of Isolation (shielding = 2) | Deaths (per 10,000) | Working Fraction | Total Deaths (per 10,000) | Working Fraction |
| 25% | 600 | 90.95% | 720 | 94.96% |
| 50% | 250 | 67.13% | 510 | 83.65% |
| 75% | 40 | 99.92% | 40 | 99.95% |

**TABLE S2:** Comparison between feedback control approach for contact policy with and without delay. Isolation efficiencies of 25%, 50% and 75% are used for the purpose of comparison, with performance metrics of total deaths and working fraction. Both of the performance metrics are nearly identical for all cases, which suggests high robustness to delay on the system.

| Feedback Control Approach | Without Delay |  | With Delay |  |
| --- | --- | --- | --- | --- |
| Efficiency of Isolation (shielding = 2) | Deaths (per 10,000) | Working Fraction | Total Deaths (per 10,000) | Working Fraction |
| 25% | 620 | 89.98% | 620 | 89.94% |
| 50% | 250 | 62.40% | 250 | 62.12% |
| 75% | 30 | 99.90% | 30 | 99.90% |

The results, comprising total death and working fraction, are presented in table S1 for various isolation efficiencies. These findings indicate higher death rates as well as higher working fractions for the delayed system except for the third, and highest efficiency of isolation, where the respective performance measures are similar. For the case of 50% isolation, the death count is more than doubled due to delay. This significant difference in performance metrics demonstrates the shortcomings of implementing a policy based on optimal control if the system is not ideal.

We also tested the sensitivities of the total death and working fraction to 30-day delays. The results, shown in table S2 below, indicate low sensitivity compared to the observed results for the optimal control solution. We compared these performance metrics obtained from applications of the optimal control vs. feedback control to the system without delay. The results, summarized in tables S1 and S2 respectively, indicate similar performance with respect to the performance metrics used. Thus, feedback control is nearly as good as optimal control if there is no delay; while having more robust features in the event that there is a delay in application.

### II. SUPPLEMENTARY TABLES

**TABLE S3:**
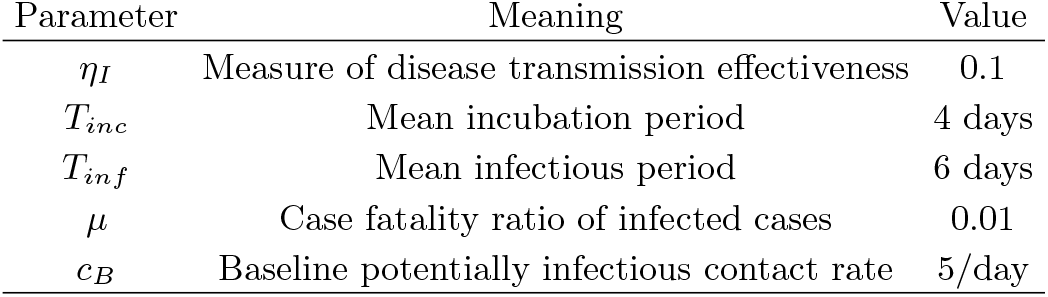
Epidemiological characteristics.

| Parameter | Meaning | Value |
| --- | --- | --- |
| $\eta_I$ | Measure of disease transmission effectiveness | 0.1 |
| $T_{inc}$ | Mean incubation period | 4 days |
| $T_{inf}$ | Mean infectious period | 6 days |
| $\mu$ | Case fatality ratio of infected cases | 0.01 |
| $c_B$ | Baseline potentially infectious contact rate | 5/day |

**TABLE S4:**
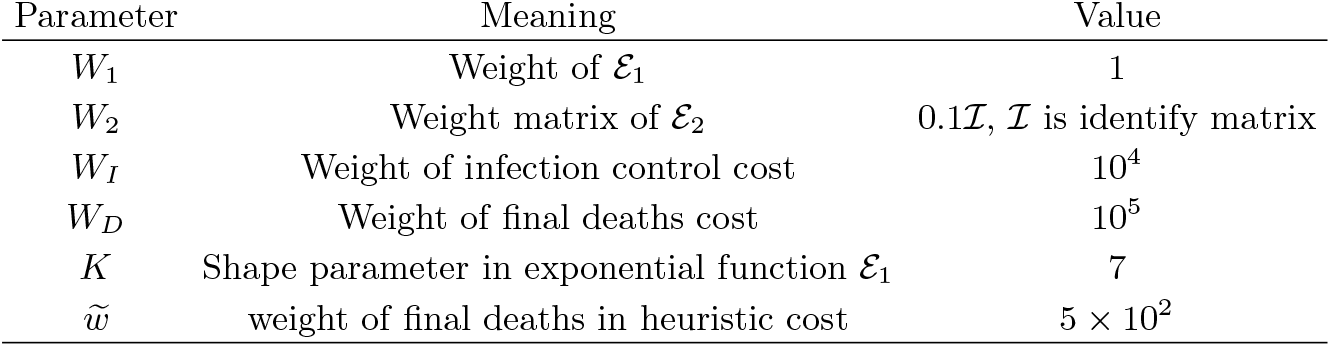
Weight regulators and optimization parameters.

| Parameter | Meaning | Value |
| --- | --- | --- |
| $W_1$ | Weight of $\mathcal{E}_1$ | 1 |
| $W_2$ | Weight matrix of $\mathcal{E}_2$ | $0.1\mathcal{I}$, $\mathcal{I}$ is identify matrix |
| $W_I$ | Weight of infection control cost | $10^4$ |
| $W_D$ | Weight of final deaths cost | $10^5$ |
| $K$ | Shape parameter in exponential function $\mathcal{E}_1$ | 7 |
| $\tilde{w}$ | weight of final deaths in heuristic cost | $5 \times 10^2$ |

---

**FIG. S4:**
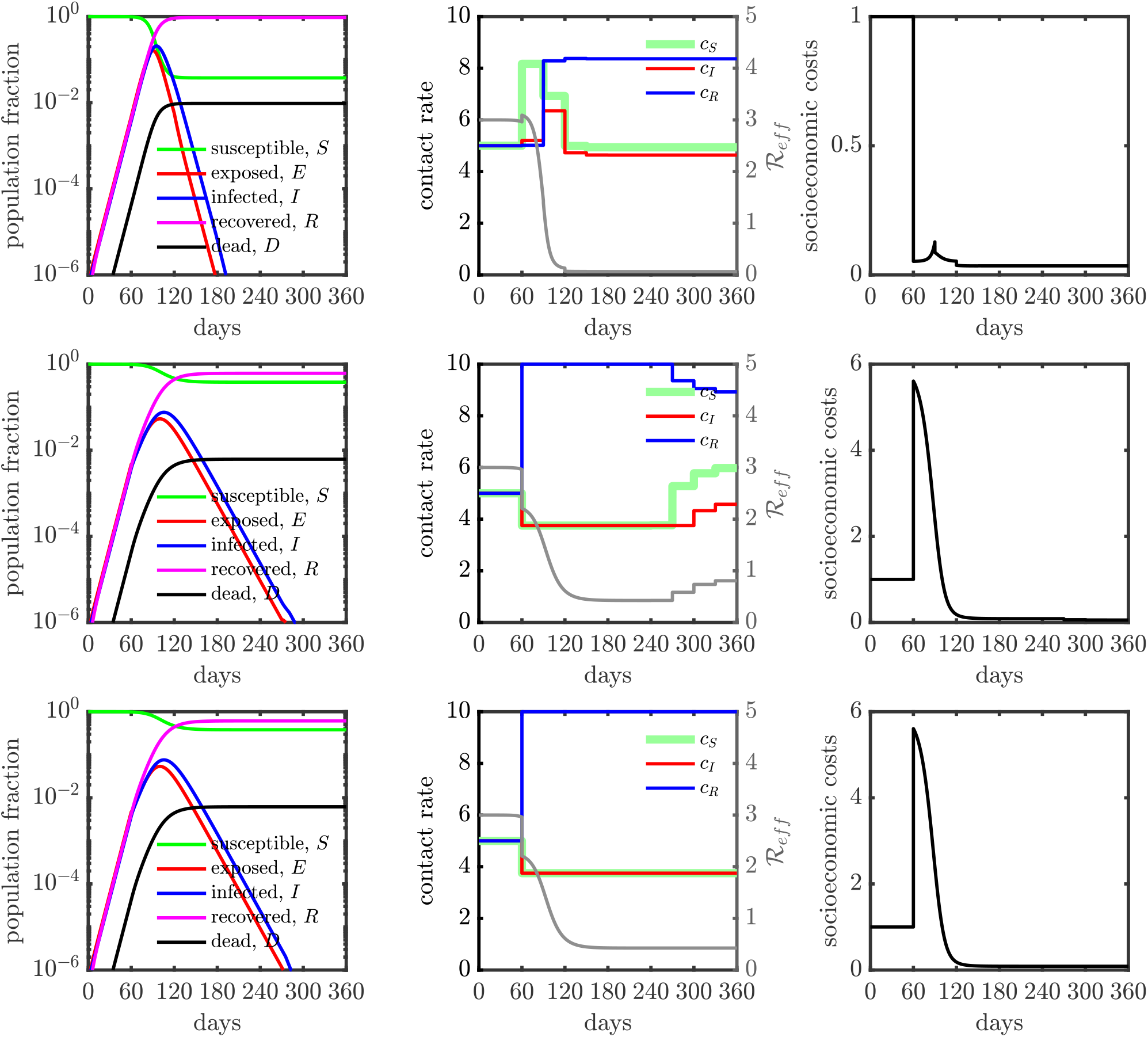
Population dynamics in a SEIR model with controlled contact rate (25% isolation efficiency). (Top) The relative importance ratio (ξ) is set as 10^−4^, the policy (*i.e*., contact rate) is deployed to primarily minimize socioeconomic costs 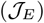. The socioeconomic costs are maintained at low level for all the policy intervention period (see the right panel after 60 days). Final state of the system in terms of population fraction is: *D*(*t_f_*) ≈ 1%, *S*(*t_f_*) ≈ 4% and *R*(*t_f_*) ≈ 95%, where *t_f_* = 360 days. The fraction of working-days recovered is about 100%. (Middle) The relative importance ratio (ξ) is set as 1, the policy (*i.e*., contact rate) is deployed to minimize infectious diseases costs 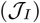 while keeping the socioeconomic impacts low. The Final state of the system in terms of population fraction is: *D*(*t_f_*) ≈ 0.6%, *S*(*t_f_*) ≈ 38% and *R*(*t_f_*) ≈ 61%, where *t_f_* = 360 days. The fraction of working-days recovered is about 91%. (Bottom) The relative importance ratio (ξ) is set as 10^4^, the policy (*i.e*., contact rate) is deployed to primarily minimize infectious diseases costs 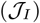. The controlled contact rates of susceptible cases (*c_S_*) and infected cases (*c_I_*) are overlapped at minimum contact rate boundary (*c_min_*). The Final state of the system in terms of population fraction is: *D*(*t_f_*) ≈ 0.6%, *S*(*t_f_*) ≈ 38% and *R*(*t_f_*) ≈ 61%, where *t_f_* = 360 days. The fraction of working-days recovered is about 88%. Here, we set *c_min_* = (3/4)*c_B_* (*i.e*., up to 25% isolation efficiency) and *c_max_* = 2*c_B_* (*i.e*., up to twice enhanced interactions).

**FIG. S5:**
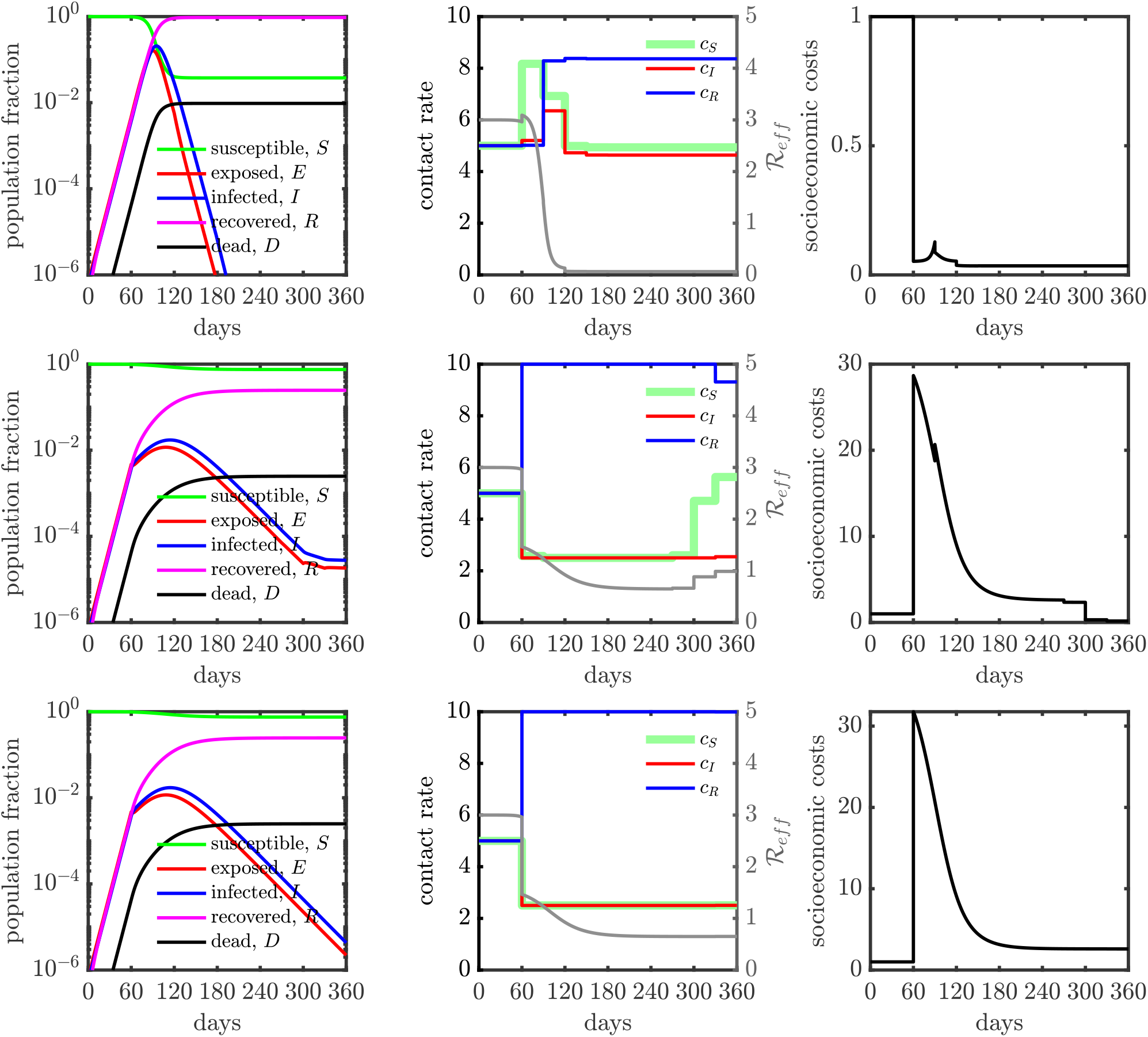
Population dynamics in a SEIR model with controlled contact rate (50% isolation efficiency). (Top) The relative importance ratio (ξ) is set as 10^−4^, the policy (*i.e*., contact rate) is deployed to primarily minimize socioeconomic costs 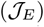. The socioeconomic costs are maintained at low level for all the policy intervention period (see the right panel after 60 days). Final state of the system in terms of population fraction is: *D*(*t_f_*) ≈ 1%, *S*(*t_f_*) ≈ 3% and *R*(*t_f_*) ≈ 96%, where *t_f_* = 360 days. The fraction of working-days recovered is about 100%. (Middle) The relative importance ratio (ξ) is set as 1, the policy (*i.e*., contact rate) is deployed to minimize infectious diseases costs 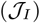 while keeping the socioeconomic impacts low. For the first half intervention period (from 60 days to 240 days), the controlled contact rates of susceptible cases (*c_S_*) and infected cases (*c_I_*) are overlapped at minimum contact rate boundary (*c_min_)*. There are huge socioeconomic impacts at beginning of policy intervention period (due to the rapid contact rate reduction of susceptible cases and infected cases, e.g. isolation), the socioeconomic impacts become lower later because of *shielding* (*i.e*., contact rate of recovered cases is enhanced). Moreover, at the last final 3 months, the socioeconomic costs drop more since the susceptible cases are back to normal (contact rate *c_S_* is increasing). The Final state of the system in terms of population fraction is: *D*(*t_f_*) ≈ 0.25%, *S*(*t_f_*) ≈ 75% and *R*(*t_f_*) ≈ 25%, where *t_f_* = 360 days. The fraction of working-days recovered is about 66%. (Bottom) The relative importance ratio (ξ) is set as 10^4^, the policy (*i.e*., contact rate) is deployed to primarily minimize infectious diseases costs 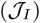. The controlled contact rates of susceptible cases (*c_S_*) and infected cases (*c_I_*) are overlapped at minimum contact rate boundary (*c_min_)* for all time. Similar to the intermediate case (ξ = 1), large socioeconomic impacts are observed at beginning of policy intervention period. The Final state of the system in terms of population fraction is: *D*(*t_f_*) ≈ 0.25%, *S*(*t_f_*) ≈ 75% and *R*(*t_f_*) ≈ 25%, where *t_f_* = 360 days. The fraction of working-days recovered is about 59%. Here, we set *c_min_* = *c_B_*/2 (*i.e*., up to 50% isolation efficiency) and *c_max_* = 2*c_B_* (*i.e*., up to twice enhanced interactions).

**FIG. S6:**
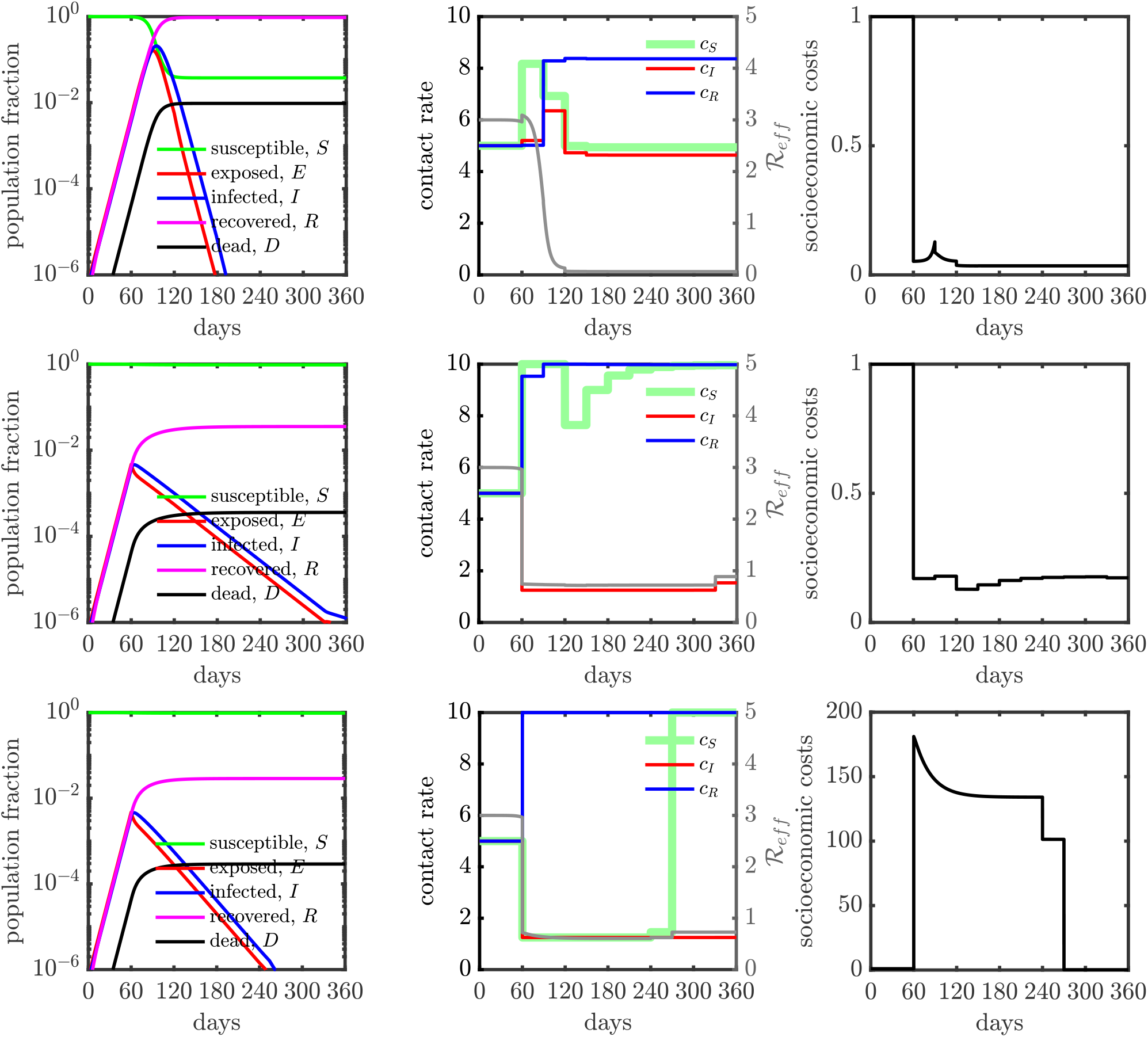
Population dynamics in a SEIR model with controlled contact rate (75% isolation efficiency). (Top) The relative importance ratio (ξ) is set as 10^−4^, the policy (*i.e*., contact rate) is deployed to primarily minimize socioeconomic costs 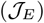. The socioeconomic costs are maintained at low level for all the policy intervention period (see the right panel after 60 days). Final state of the system in terms of population fraction is: *D*(*t_f_*) ≈ 1%, *S*(*t_f_*) ≈ 4% and *R*(*t_f_*) ≈ 95%, where *t_f_* = 360 days. The fraction of working-days recovered is about 100%. (Middle) The relative importance ratio (ξ) is set as 1, the policy (*i.e*., contact rate) is deployed to minimize infectious diseases costs 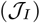 while keeping the socioeconomic impacts low. Infected individuals are locked down at home for all intervention period while susceptible and recovered individuals are free to go out. The Final state of the system in terms of population fraction is: *D*(*t_f_*) ≈ 0.04%, *S*(*t_f_*) ≈ 96% and *R*(*t_f_*) ≈ 4%, where *t_f_* = 360 days. The fraction of working-days recovered is about 100%. (Bottom) The relative importance ratio (ξ) is set as 10^4^, the policy (*i.e*., contact rate) is deployed to primarily minimize infectious diseases costs 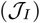. The controlled contact rates of susceptible cases (*c_S_*) and infected cases (*c_I_*) are overlapped at minimum contact rate boundary (*c_min_*) at beginning, then the susceptibles are free to move later. The Final state of the system in terms of population fraction is: *D*(*t_f_*) ≈ 0.03%, *S*(*t_f_*) ≈ 97% and *R*(*t_f_*) ≈ 3%, where *t_f_* = 360 days. The fraction of working-days recovered is about 50%. Here, we set *c_min_* = *c_B_*/4 (*i.e*., up to 75% isolation efficiency) and *c_max_* = 2*c_B_* (*i.e*., up to twice enhanced interactions).

**FIG. S7:**
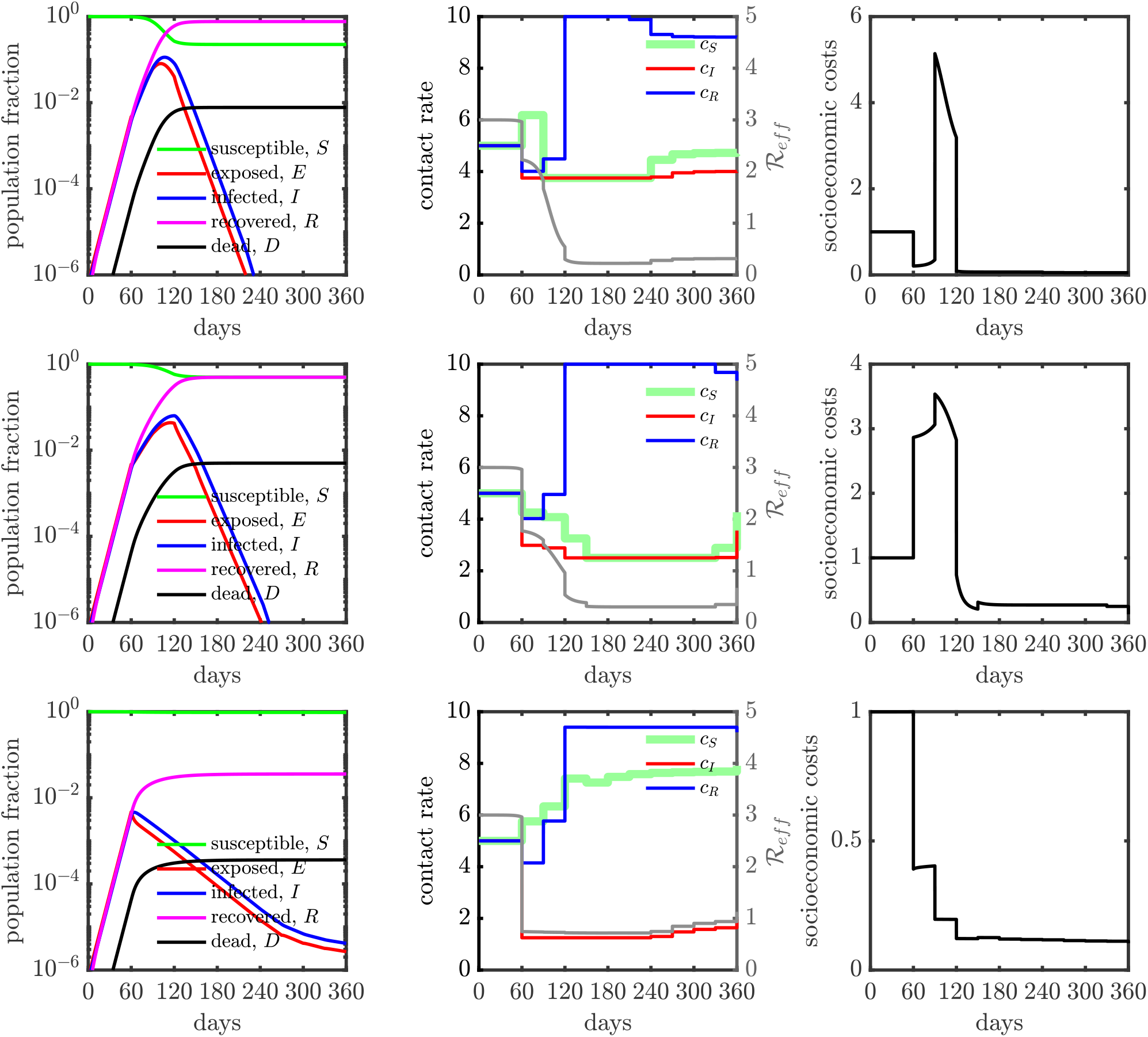
Population dynamics in a SEIR model with mis-timed control policy for various isolation efficiencies: (Top) 25% isolation efficiency; (Middle) 50% isolation efficiency and (Bottom) 75% isolation efficiency. The relative importance (ξ) is 1 for all the cases. The optimal control policy is computed for a system one month after an outbreak. Then, we enforce the optimal control policy 30 days later, *i.e*., at the end of 60 days after the start of the outbreak. The contact rate interventions start at 60 days. The total deaths and working fraction for the delayed system are presented in table S1 for various isolation efficiencies. The optimal control policies and their corresponding dynamics without mistiming are shown in the middle row of Figs. S4, S5, S6.

**FIG. S8:**
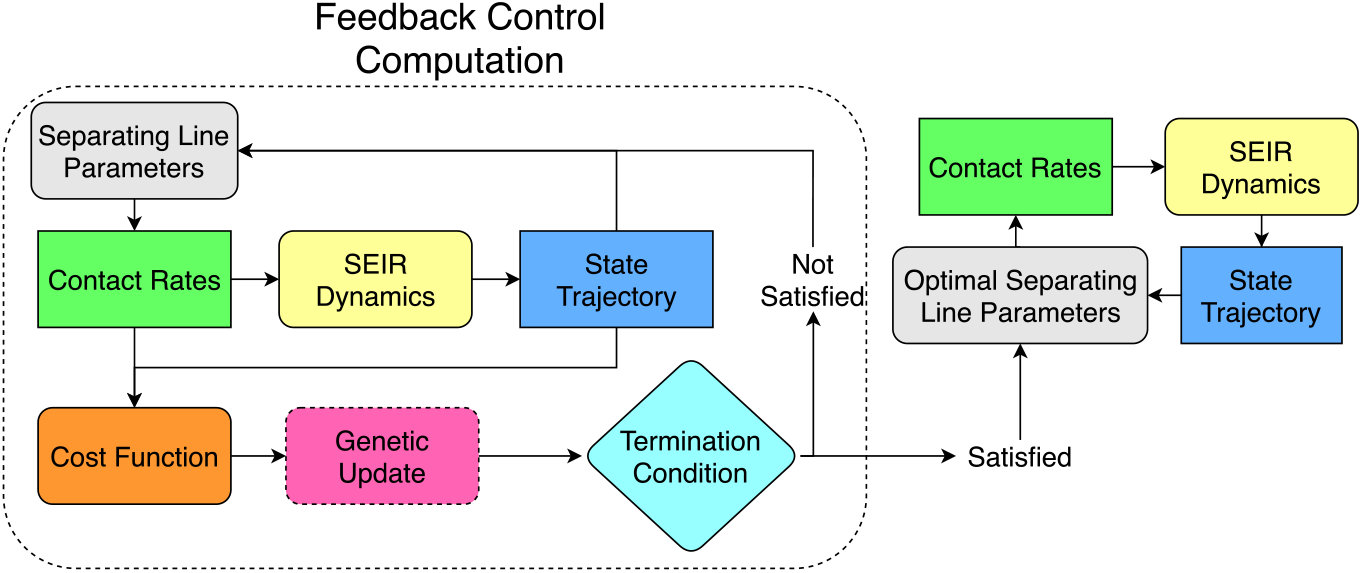
Flowchart for computing state dependent heuristic policy. The line dividing the *I-R* plane is parameterized and for a particular parameter set, the contact rate is determined solely from the current infected and recovered cases. Starting with an initial parameter guess, the contact rate is computed from the current *I* and *R* values, giving the system’s state at the next time instant. After the full state trajectory is computed, the associated cost is calculated and a genetic update step is taken which identifies the next parameters. If the termination condition is satisfied, these parameters are passed on as the optimal parameters which can be used to compute the optimal contact policy. If the termination condition is not satisfied, the line parameters are updated and the loop is run again.

**FIG. S9:**
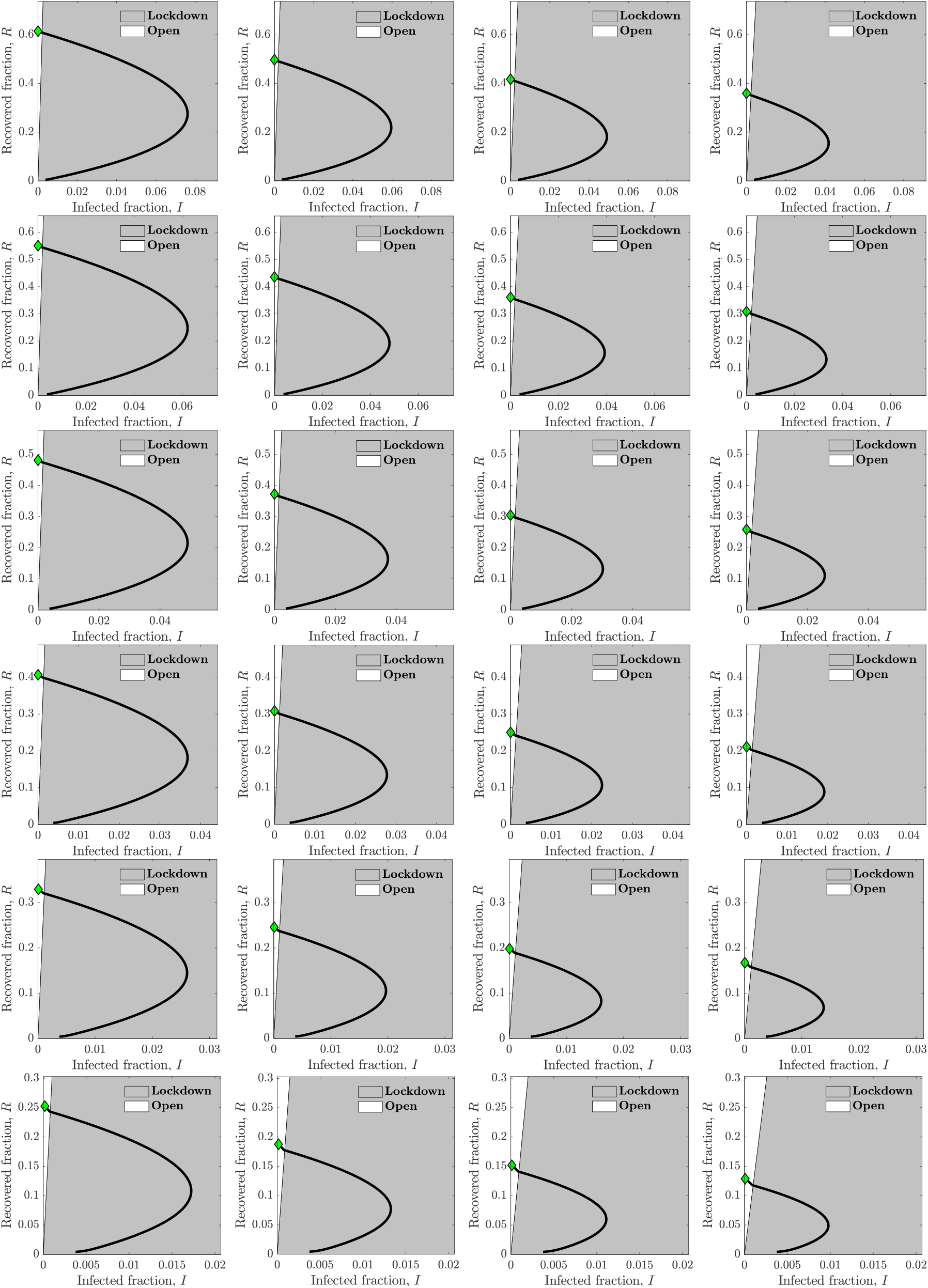
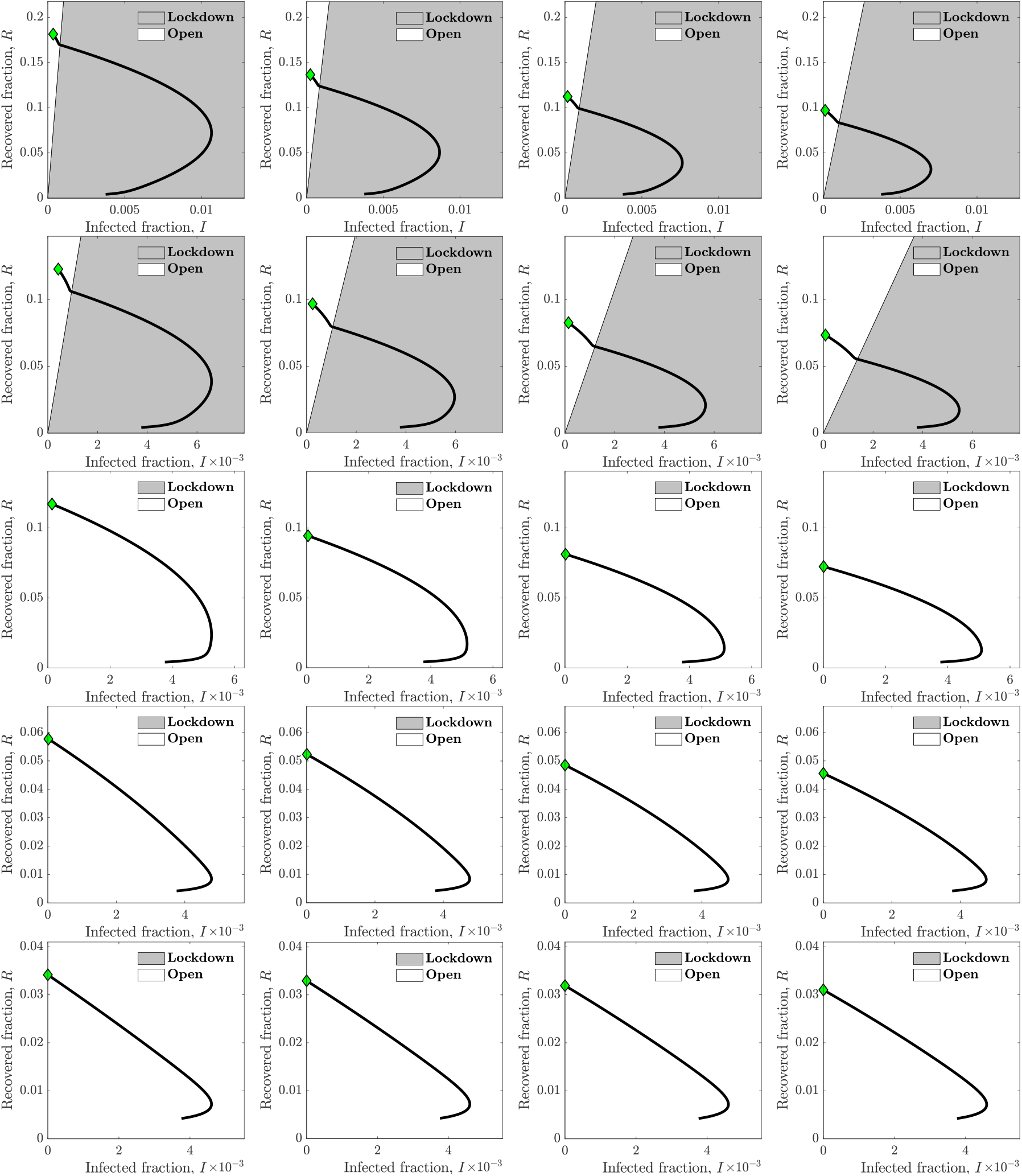
Heuristic state feedback intervention policies varying with isolation efficiency and shielding level on *I — R* phase plane. The isolation efficiency for the first row is 25% and each row increases the efficiency by 5%, which gives the efficiency of the last row as 75%. Next, the 4 columns correspond to shielding levels of 2, 3, 4 and 5 from left to right. In each plot, the grey region specifies the area of the *I* — *R* plane where a lockdown policy is enforced for the susceptible population and the white region gives the area where the lockdown is lifted i.e. the system is ’open’. The phase trajectory (*I*(*t*), *R*(*t*)), *i.e*., the black line, crosses from the lockdown to open region when it intersects the dividing line and this crossover happens at most once. The final state of the system is marked by a green diamond.

## Notes

### Competing Interest Statement

The authors have declared no competing interest.

### Funding Statement

We thank Weitz Group team members for comments and feedback. TThis work was supported by grants from the Army Research Office (W911NF1910384), National Institutes of Health (1R01AI46592-01), and the National Science Foundation (1806606 and 2032082).

### Summary of Updates

All the pages fit in a 8.5 x 11 inches sheet.

